# Plasma gradient of soluble urokinase-type plasminogen activator receptor is linked to pathogenic plasma proteome and immune transcriptome and stratifies outcomes in severe COVID-19

**DOI:** 10.1101/2021.06.19.21259125

**Authors:** Jafar Sarif, Deblina Raychaudhuri, Ranit D’Rozario, Purbita Bandopadhyay, Praveen Singh, Priyanka Mehta, Md. Asmaul Hoque, Bishnu Prasad Sinha, Manoj Kushwaha, Shweta Sahni, Priti Devi, Partha Chattopadhyay, Shekhar Ranjan Paul, Yogiraj Ray, Kausik Chaudhuri, Sayantan Banerjee, Debajyoti Majumdar, Bibhuti Saha, Biswanath Sharma Sarkar, Prasun Bhattacharya, Shilpak Chatterjee, Sandip Paul, Pramit Ghosh, Rajesh Pandey, Shantanu Sengupta, Dipyaman Ganguly

## Abstract

Disease caused by SARS-CoV-2 coronavirus (COVID-19) has resulted in significant morbidity and mortality world-wide. A systemic hyper-inflammation characterizes the severe COVID-19 disease often associated with acute respiratory distress syndrome (ARDS). Blood biomarkers capable of risk stratification are of great importance in effective triage and critical care of severe COVID-19 patients. In the present study we report higher plasma abundance of soluble urokinase-type plasminogen activator receptor (sUPAR), expressed by an abnormally expanded circulating myeloid cell population, in severe COVID-19 patients with ARDS. Plasma sUPAR level was found to be linked to a characteristic proteomic signature of plasma, linked to coagulation disorders and complement activation. Receiver operator characteristics curve analysis identified a cut-off value of sUPAR at 1996.809 pg/ml that could predict survival in our cohort (Odds ratio: 2.9286, 95% confidence interval 1.0427-8.2257). Lower sUPAR level than this threshold concentration was associated with a differential expression of the immune transcriptome as well as favourable clinical outcomes, both in terms of survival benefit (Hazard ratio: 0.3615, 95% confidence interval 0.1433-0.912) and faster disease remission in our patient cohort. Thus we identified sUPAR as a key pathogenic circulating molecule linking systemic hyperinflammation to the hypercoagulable state and stratifying clinical outcomes in severe COVID-19 patients with ARDS.

## INTRODUCTION

The ongoing pandemic caused by the severe acute respiratory syndrome causing coronavirus 2 (SARS-COV-2) has resulted in close to 175 million documented infections and more than 3.7 million deaths. The respiratory disease caused by SARS-COV-2 or COVID-19 at times progresses to acute respiratory distress syndrome (ARDS), often with fatal outcomes in some patients [1]. Severe COVID-19 is characterized by a hyper-inflammation, the key manifestations being a systemic cytokine deluge and an abnormal myeloid expansion among circulating immune cells [2-5]. In addition to the hyper-inflammation, a number of patients with severe COVID-19 also present with intravascular coagulation as well as abnormal complement activation [6-9]. Thus the systemic cytokine surge, a hyper-coagulable state and tissue damage mediated by complement activation are the three established pathogenic triads in these patients. These also offer the possibility of identifying circulating factors with predictive potential for disease outcomes. Risk-stratifying biomarkers, which can be probed early enough in severe COVID-19 patients, can be useful as pre-assessors for effective triage or intensive care in low-resource settings.

Previous single cell RNA sequencing (scRNAseq) studies in severe COVID-19 revealed abnormally expanded circulating myeloid cell compartment as well as an enriched expression of urokinase type plasminogen activator receptor (PLAUR gene, expressing UPAR protein) [3,4]. UPAR (or CD87) is a glycosylphosphatidylinositol (GPI)-anchored receptor present on the surface of various cells, including immune cells including monocytes, macrophages and neutrophils. Cell surface UPAR binds UPA, and transforms plasminogen into plasmin, which in turn initiates a proteolytic cascade to degrade the components of the ECM [10]. UPAR also lies within the complex regulatory network of the coagulation cascade as well as complement activation and complement mediated pathogen or host cell clearance [11-13].

Increased plasma abundance of soluble UPAR has been documented widely in chronic inflammatory contexts [6], thus it is also proposed to be a potential pathogenic molecule involved in the acute systemic hyper-inflammation in COVID-19 [10, 14]. In a cohort of COVID-19 patients from India, which was originally recruited for a randomised control trial on convalescent plasma therapy, we found significantly high plasma levels of soluble UPAR in severe COVID-19 patients early in the course of severity, which correlated with an expanded myeloid cell compartment in circulation. A characteristic proteomics signature of activation of coagulation cascade as well as complement activation was found to be associated with higher plasma concentrations of UPAR, as were specific immune related pathways enriched in peripheral blood transcriptome analysis. Finally we found that patients, suffering from ARDS in severe COVID-19 but with plasma levels of sUPAR below a computed cut-off value, registered significantly more favourable disease outcomes.

## RESULTS

### Expansion of circulating CD11c ^+^ HLA-DR^—^ myeloid cells expressing sUPAR in severe COVID-19 patients

COVID-19 patients with mild symptoms (N=8, Age=41.5±18.95 years) or ARDS (N=77, Age =61±11.86 years) were recruited at the ID & BG Hospital, Kolkata, India (Supplemental table 1). Peripheral blood sampling was done on the day of enrolment with due ethical approval from the institutional review board. Frequency of circulating CD11c^+^HLA-DR^--^ myeloid cells was assessed by flow cytometry. On comparing relative abundance of circulating CD11c^+^HLA-DR^---^ proinflammatory macrophages between the COVID-19 patients with mild diseases and patients who progressed to ARDS, we found a prominent expansion of these cells in patients suffering from ARDS (Figure 1A, B), as has been reported by a number of previous studies [4-6]. We found no correlation between the abundance of these cells in circulation in ARDS patients and the plasma levels of most of the cytokines (among the 36 selected based on detectable levels in at least 70% of the ARDS patients in the cohort, data not shown), except for Eotaxin (Pearson R=0.3725, P=0.0029). HGF (Pearson R=0.2503, P=0.0497) and IL-1α (Pearson R=0.2588, P=0.0423) (Figure 1C-E).

**Fig. 1.**
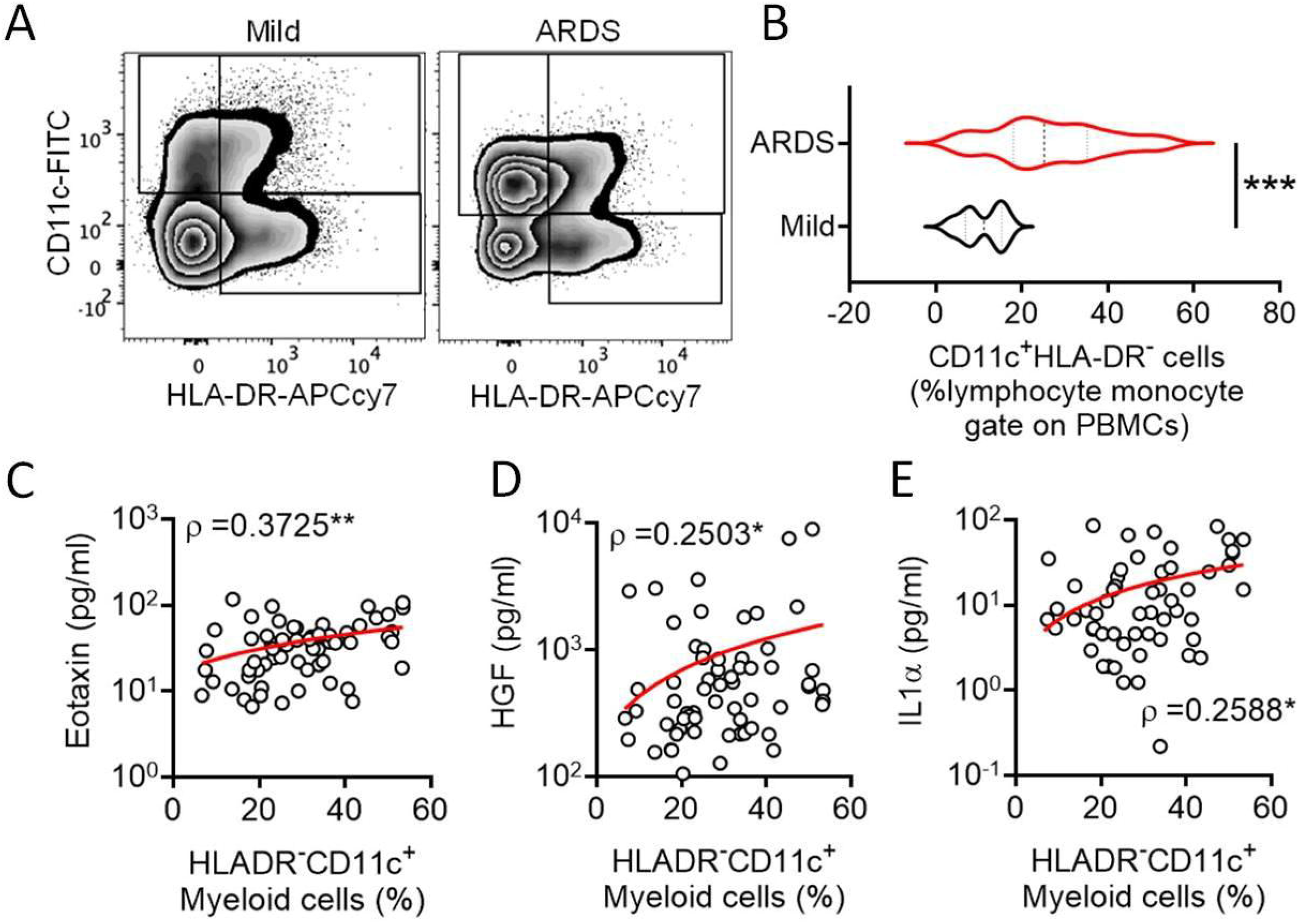
Expansion of circulating CD11c^+^HLA-DR^-^ myeloid cell subset in severe COVID-19. (A) Representative flow cytometry plots for gating of circulating CD11c^+^HLA-DR^-^ myeloid cells from COVID-19 patients with either mild disease or ARDS. (B) Violin plots showing frequency of circulating CD11c^+^HLA-DR^-^myeloid cells compared between COVID-19 patients with either mild disease or ARDS. Mann Whitney test was performed. (C) - (E) Correlation between the frequency of circulating CD11c^+^HLA-DR^-^myeloid cells and plasma level of the cytokines Eotaxin (C), HGF (D) and IL-1α (E) is plotted. Spearman ρ values are shown, **P <0.005, *P<0.05.

Interestingly, a recent scRNAseq study reported an enriched expression of PLAUR, the gene for UPAR, in the similarly expanded myeloid cell compartment in severe COVID-19 [3]. It intrigued us to explore if the circulating CD11c^+^HLA-DR^-^myeloid cells in severe COVID-19 patients are enriched for UPAR expression. To this end we analyzed two public datasets on scRNAseq, one done on whole blood (GSE163668, Figure 2A, B) and the other on cells in the bronchoalveolar lavage fluid (GSE145926, Figure 2C, D) from COVID-19 patients with different disease states [15, 16]. We found that the CD11c^high^HLA-DR^low^ myeloid cell subsets were highly abundant in patients with severe COVID-19 both in the circulation as well as in the airways on these analyses as well as noted a highly enriched expression of PLAUR in these CD11c^high^HLA-DR^low^ myeloid cells (Figure 2B, D).

**Fig. 2.**
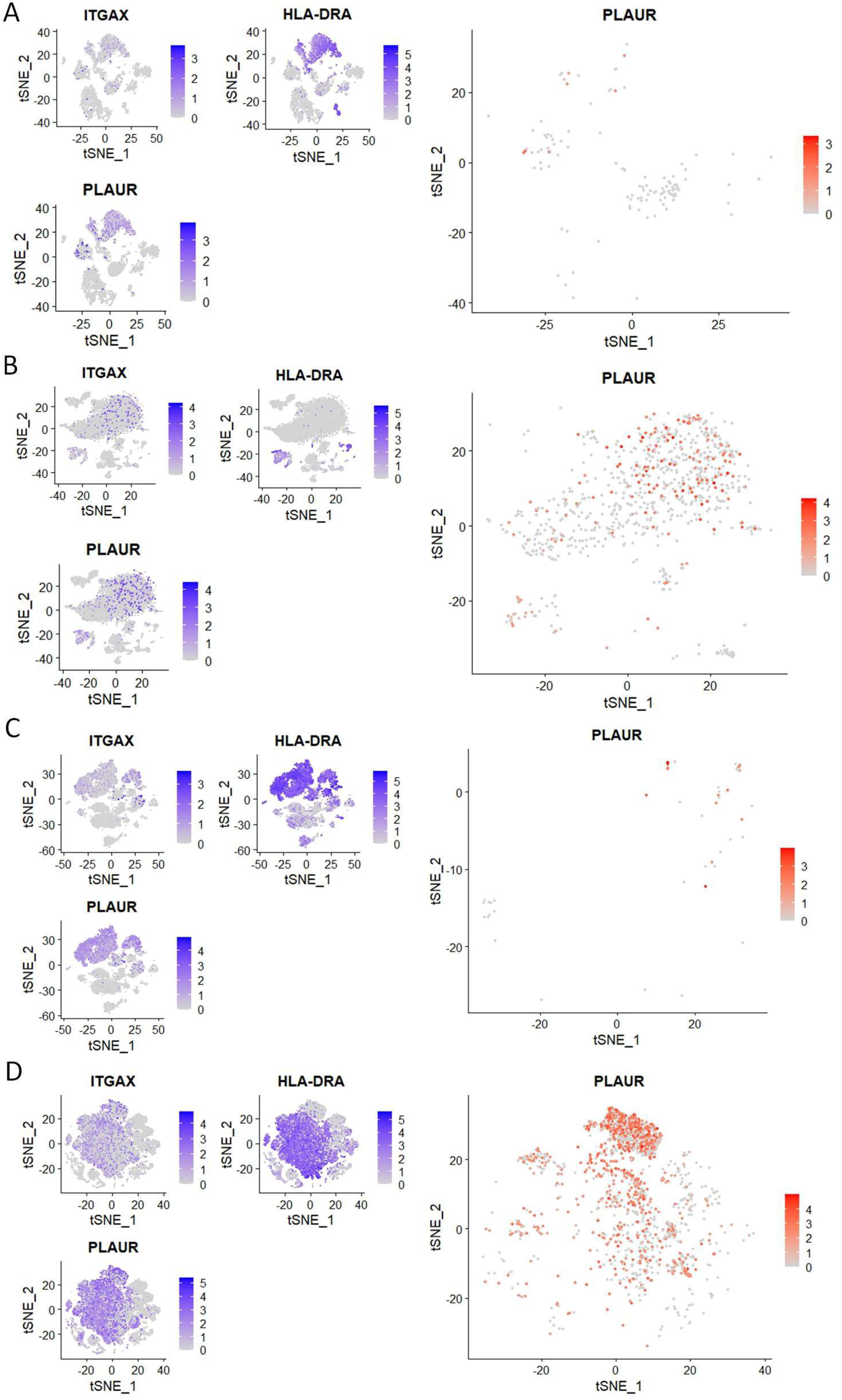
Reanalysis of single cell sequencing data to confirm myeloid sourcing of sUPAR. (A) and (B) Analysis of scRNAseq data (GSE163668) to compare frequency of peripheral blood CD11c^high^HLA-DR^low^cells and expression of PLAUR in them between patients with mild (left panel) or severe (right panel) COVID-19 disease. (C) and (D) Analysis of scRNAseq data (GSE GSE145926) to compare frequency of CD11c^high^HLA-DR^low^cells in the bronchoalveolar lavage fluid and expression of PLAUR in them between patients with mild (left panel) or severe (right panel) COVID-19 disease. In both cases, three smaller plots on the left show distribution of expression of ITGAX (gene for CD11c), HLA-DRA (gene for HLA-DR) and PLAUR (gene for UPAR) among all cells. The bigger plot on the right shows expression of PLAUR among CD11c^high^HLA-DR^low^cells.

### Higher plasma abundance of sUPAR in severe COVID-19 is linked to systemic cytokine surge and TNFα-activated myeloid cells

Next we measured the level of sUPAR in plasma samples from the patients in our cohort and found a significant correlation between the abundance of the circulating CD11c^+^HLA-DR^-^ myeloid cells and plasma sUPAR levels (Figure 3A). Moreover, a significantly higher abundance of sUPAR was noted in patients who have progressed to ARDS, compared to patients with milder symptoms (Figure 3B). The plasma level of sUPAR had no relationship with either the viral load of the ARDS patients at the time of plasma sampling (Figure 3C) or the neutralizing antibody content of their plasma (Figure 3D). Age or gender of the patients also did not show any relationship with the plasma levels of sUPAR (Figure 3 E, F).

**Fig. 3.**
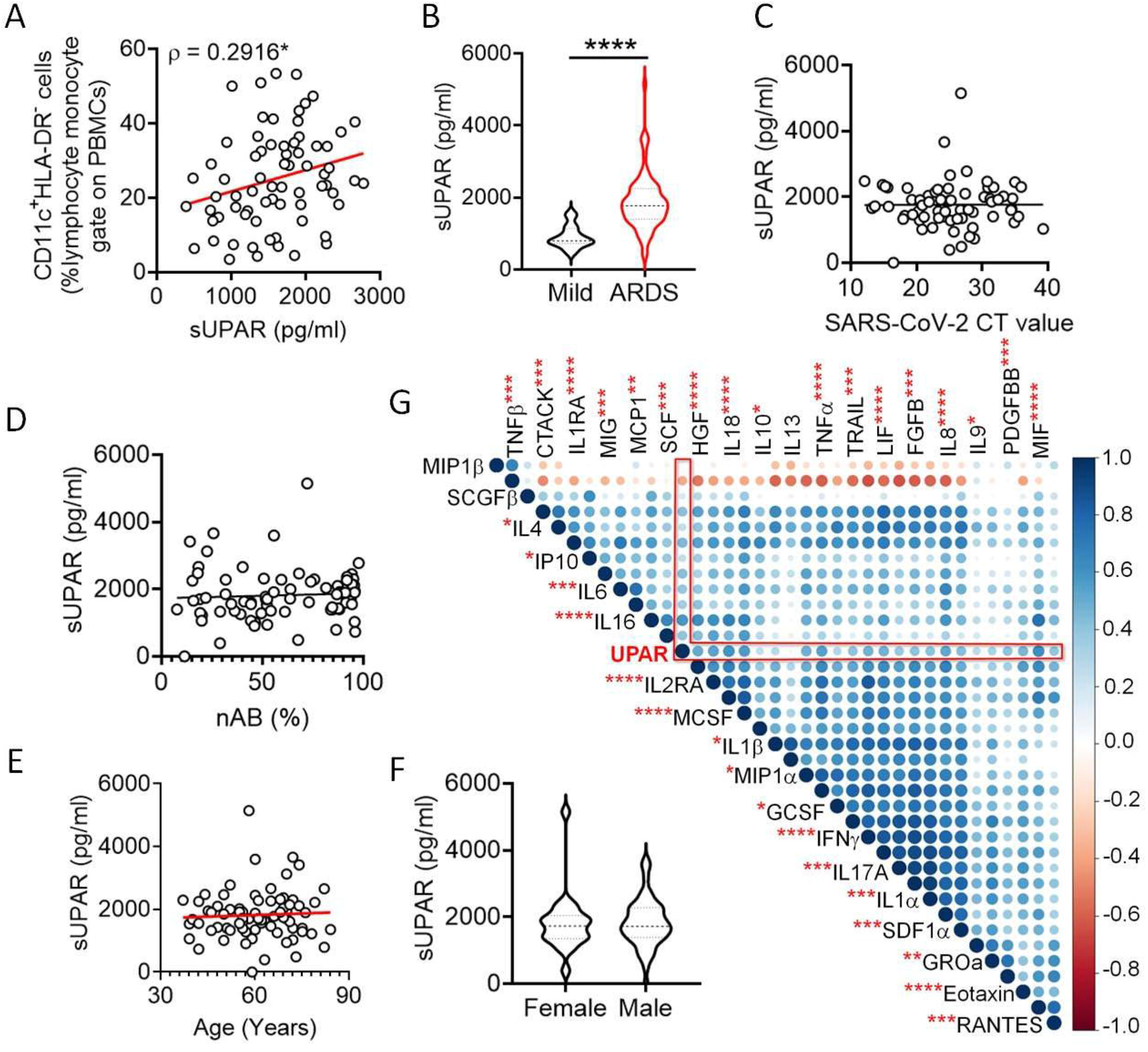
Increased plasma abundance of soluble UPAR and its cellular source in severe COVID-19. (A) Correlation between frequency of circulating CD11c^+^HLA-DR^--^myeloid cells and plasma levels of sUPAR in COVID-19 patients. (B) Plasma level of sUPAR compared between COVID-19 patients with either mild disease or ARDS. (C) No correlation between average CT values for two SARS-CoV-2 genes and the plasma concentration of sUPAR. (D) No correlation between percent neutralization function of patient plasma (derived from a surrogate assay measuring neutralization of receptor binding protein and ACE2 binding) and the plasma concentration of sUPAR. (E) No correlation between age of patients with ARDS and the plasma level of sUPAR. (F) Comparison between male and female patients with ARDS for the plasma level of sUPAR. Mann Whitney test was performed. (G) Corr plot showing mutual correlation between the plasma levels of 36 cytokines and soluble UPAR in COVID-19 patients with ARDS. The dots are color-coded and size-scaled for the Spearman ρ values for individual correlations, significance levels for correlation between plasma levels of sUPAR and individual cytokines are noted as superscripts on cytokine names, ****P<0.0001, ***<P0.0005, **P<0.005, *P<0.05.

Plasma level of sUPAR was significantly correlated with plasma abundance of individual entities (in total 36 different cytokines) making up the systemic cytokine deluge in the ARDS patients (Figure 3G). This wide correlation of the plasma abundance of the cytokines with that of sUPAR may mechanistically represent the effect of some of the proinflammatory cytokines on the expanded myeloid cells, that leads to expression of sUPAR. On the other hand, it also presumably represents the amplified systemic inflammatory circuit leading to a correlated abundance. We noted a prominently significant correlation of plasma sUPAR concentration with plasma level of tumor necrosis alpha (TNFα), a major inflammatory cytokine capable of activating myeloid cells. In a recent study monocyte-derived macrophages were stimulated with different cytokines and scRNAseq was performed to discover the heterogeneity of transcriptional responses [17]. To shed some light on the mechanistic aspects of myeloid cell expression of sUPAR, we analyzed this data retrieved from GEO Datasets (GSE168710) with an aim to look for expression of PLAUR in these macrophages as they respond to different cytokine stimuli (Figure 4A, B). While expression of PLAUR was noted in almost all the clusters of stimulated macrophages in this study (Figure 4C), stimulation with TNFα in the absence of type I or type II interferons drove the macrophages to the highest expression of PLAUR (Figure 4D). The CD11c^high^HLA-DR^low^ myeloid cells had a very high expression of PLAUR (Figure 4E), and again the highest expression was in response to TNFα (Figure 4F). Thus TNFα, with its circulating levels ramped up in the context of the systemic cytokine deluge may play a key role in inducing sUPAR expression in the abnormally expanded CD11c^high^HLA-DR^low^ myeloid cells in the severe ARDS patients, though this warrants further mechanistic exploration.

**Fig. 4.**
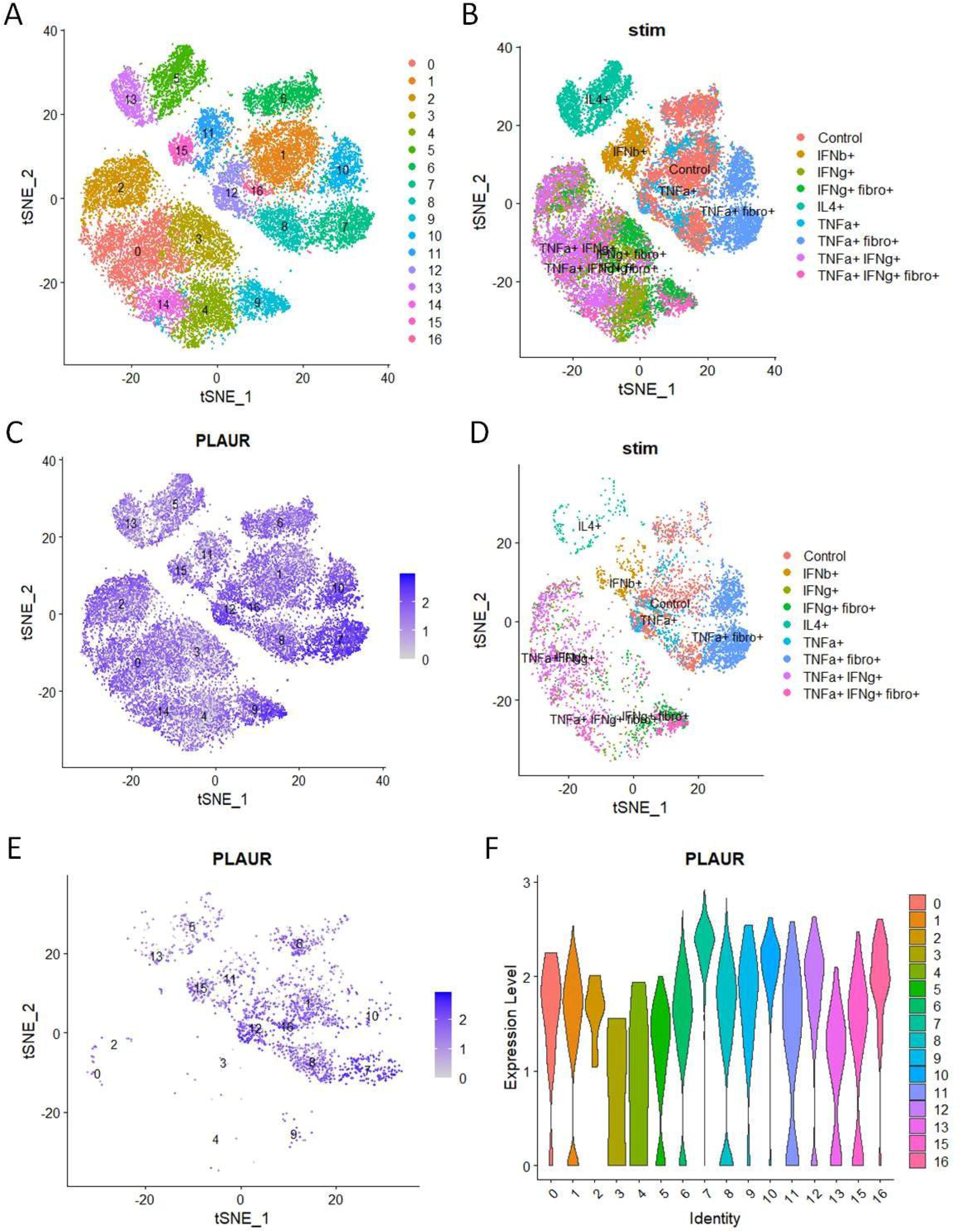
Reanalysis of single cell sequencing data to confirm role of TNFa on myeloid expression of sUPAR. (A) TSNE plot of scRNA-seq data (obtained from GSE168710) of macrophages grouped into various clusters based on their gene expression profiles. (B) TSNE plot of the same scRNA-seq data showing the different treatments to which the macrophages were exposed before sequencing. (C) TSNE plot showing the distribution of expression of the ‘PLAUR’ gene in all the cells of the same dataset. (D) TSNE plot depicting the treatment conditions to which all the PLAUR^high^ cells belong. (E) TSNE plot depicting the expression pattern of ‘PLAUR’ specifically in CD11c^high^HLA-DR^low^ cells. (F) Violin plot highlighting the difference in expression of ‘PLAUR’ between the different clusters of CD11c^high^HLA-DR^low^cells.

### Linking plasma sUPAR abundance and inflammatory plasma proteome in severe COVID-19

As discussed earlier, sUPAR is functionally involved in the intricate regulation of both the coagulation cascade and complement activation. Thus to glean further insights on the role of sUPAR in the pathogenesis of severe COVID-19 disease a proteomics analysis of the plasma was more insightful. We selected plasma samples across the range of different sUPAR values and did a proteomics analysis (Figure 5A). We could identify 179 proteins in our mass spectrometry based study (Supplemental table 2). The area under curve for the m/z values of the respective peaks were used for a semi-quantitative analysis of the abundance of those proteins in circulation. Plasma abundance of twenty-four proteins showed statistically significant correlation with the plasma level of sUPAR in the same plasma samples (Figure 5B). Significant correlative clustering of these proteins were also apparent to a variable extents. When we looked closely, abundance of a number of these proteins were showing a nice gradient, some increasing, while the others were decreasing, with increasing sUPAR concentrations in plasma (Figure 5C, supplemental table 3).

**Fig. 5.**
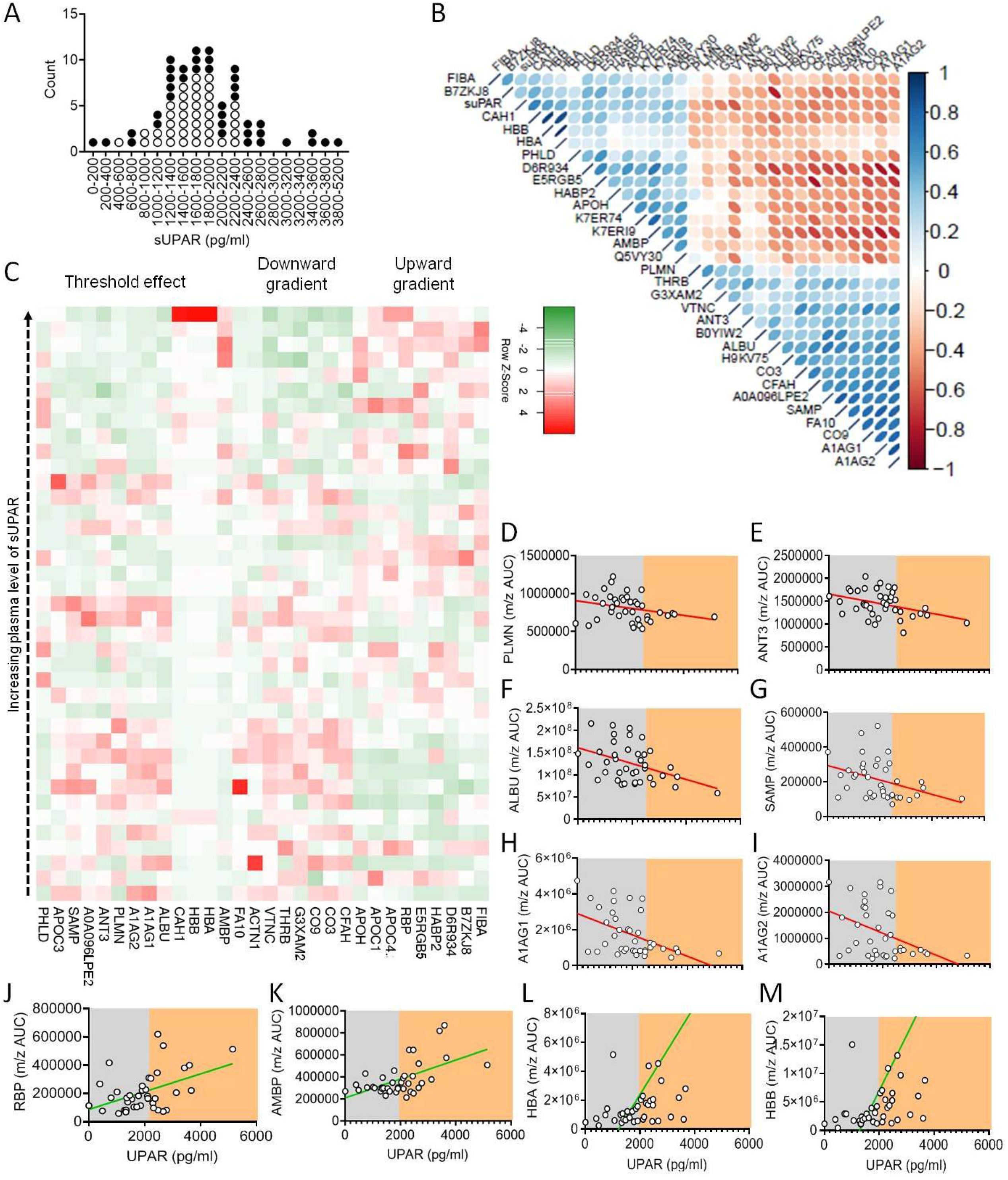
Proteomics analysis of plasma from patients with different plasma levels of sUPAR. (A) Plot showing distribution of patients and their selection for proteomic studies based on their plasma concentration of sUPAR. Each circle represents an individual patient and the filled circles represent the samples selected for mass-spectrometry based proteomic studies. Patients were randomly selected from distinct ranges of plasma sUPAR values (total of 40 patients) as indicated, with the aim of being able to represent most of the ranges. (B) Corrplot depicting two-tailed Spearman correlation coefficients between the quantities of indicated proteins as well as sUPAR in plasma of Covid-19 patients, generated using the ‘Hmisc’ package in R. (C) Heatmap depicting the relation between the concentration of sUPAR and the quantities of selected proteins (those which show significant correlations indicated, in plasma of Covid-19 patients. Each square represents one protein of one specific patient. The proteins have been categorised into three groups based on their pattern of expression-those which show a gradual increase with increase in sUPAR, those which show a gradual decrease with increase in sUPAR and those which show sudden but significant change at a threshold plasma concentration of sUPAR. (D-M) Representative line graphs for correlation between plasma sUPAR concentration and area under curve of m/z peaks of proteins PLMN (D), ANT3 (E), ALBU (F), SAMP (G), A1AG1 (H), A1AG2 (I), RBP (J), AMBP (K), HBA (L) and HBA (M), to demonstrate the threshold effect, shown by arbitrary shaded demarcations.

The identities and the functions of these proteins allowed us to appreciate a prominent signature of increased intravascular coagulation and complement activation (Supplemental table 3). For example increasing alpha fibrinogen (FIBA), hyaluronan binding protein 2 (HABP2), and decreased abundance of plasminogen (PLMN), thrombin (THRB), factor X (FA10), antithrombin III (ANT3), and vitronectin (VTNC) points to systemic hyper-coagulation associated with higher plasma sUPAR levels (Figure 5C, supplemental table 3). On the other hand, complement C1q subunit B (D6R934) showing a positive correlation and complement factor 3 (CO3), vitronectin (VTNC), complement component 9 (CO9), complement factor H (CFAH) and complement factor I (G3XAM2) showing a negative correlation with plasma sUPAR abundance, point to an increased complement activation state in patients with higher sUPAR levels (Figure 5C, supplemental table 3).

Interestingly, we noted a prominent threshold state change for a number of proteins at a particular level of plasma sUPAR abundance (Figure 5C). Notable among them were decrease in a few established anti-inflammatory acute phase reactants like anti-coagulation factors like PLMN, ANT3, serum amyloid proteins (viz. SAMP), albumin (ALBU) and the alpha 1 acid glycoproteins (A1AG1 and A1AG2) and an increase in carbonic anhydrase 1 (CAH1), a metabolic enzyme known to have proinflammatory effect on myeloid cells, retinol binding protein (RBP) shown to play a role in inflammatory endotheliopathy and haemoglobin alpha and beta chains (HBA and HBB) (Figure 5D-M).

### Identification of a pro-survival cut-off for plasma level of sUPAR linked to distinct immune transcriptome

We were intrigued by the state-change pattern of these proteins at a certain threshold level of plasma sUPAR level and wanted to explore if a cut-off value of plasma sUPAR concentration can be of interest in terms of predictive value towards clinical outcomes of the severe COVID-19 patients. We performed a ROC curve analysis for prediction of fatal outcomes of the disease. ROC analysis derived a cut-off value of 1996.809 pg/ml (Figure 6A). Although sensitivity (54.55%) and specificity (74.55%) as well as area under curve (0.619) on this analysis was not commensurate for sUPAR being a prognostic biomarker, in a proportional odds analysis too, patients with sUPAR levels higher than the cut-off value had significantly higher odds ratio for meeting with fatal outcomes (Figure 6B).

**Fig. 6.**
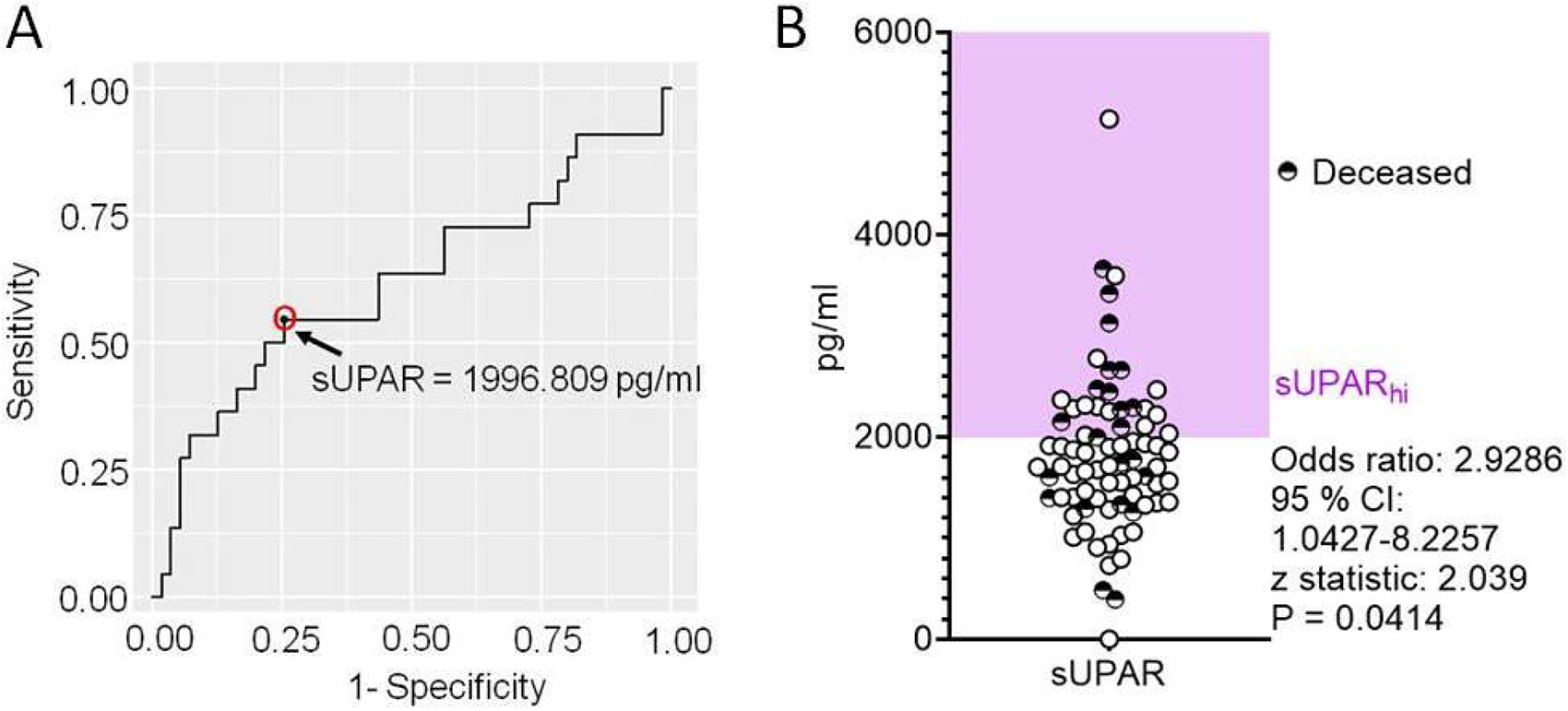
Deriving a cut-off for plasma level of soluble UPAR linked to disease outcomes in severe COVID-19. (A) Receiver operator characteristics curve for plasma levels of sUPAR as a predictor for fatal outcomes in severe COVID-19 patients. (B) Scatter plot showing plasma level of sUPAR for individual patients also marking their final disease outcomes. Odds ratio were calculated for sUPAR_hi_ patients to meet with fatal outcomes.

To further explore if this cut-off value of plasma level of sUPAR is mechanistically linked to gene expression patterns in the circulating immune cells we performed total RNA sequencing of peripheral blood cells. We selected nine representative patients for this analysis, three having low sUPAR levels, three with plasma sUPAR levels just below the cut-off value and another three from patients with plasma sUPAR values higher than the cut-off (Figure 7A).

**Fig. 7.**
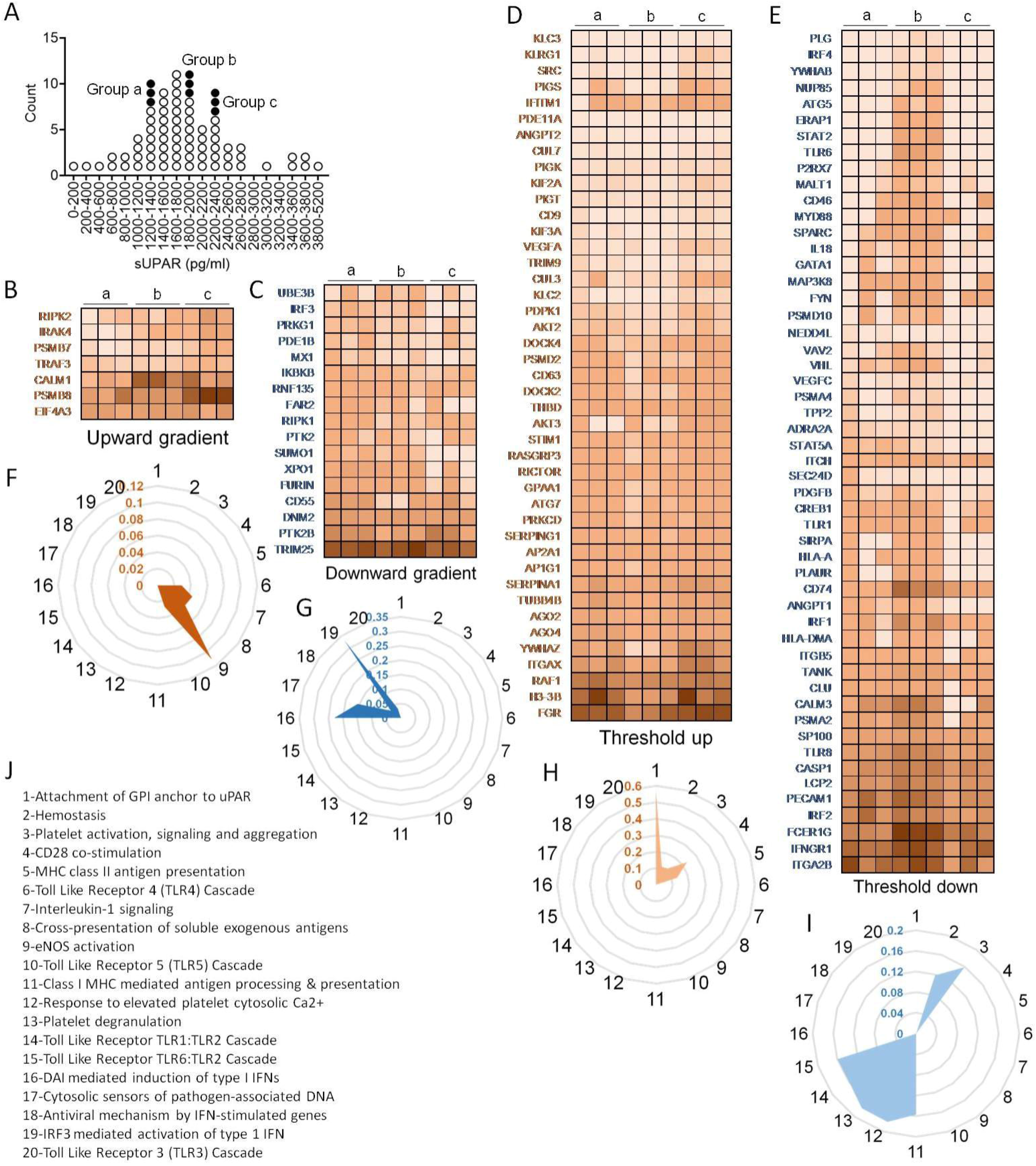
Peripheral blood transcriptome analysis of representative patients. (A) Plot showing distribution of patients and their selection for RNA sequencing based on their plasma concentration of sUPAR. Each circle represents an individual patient and the filled circles represent the samples selected for RNA sequencing. Three patients each were randomly selected from three distinct ranges of plasma sUPAR values (3 each in groups ‘a’, ‘b’ and ‘c’, total of 9 patients) as indicated. Group ‘a’ patients had low sUPAR plasma levels, the group ‘b’ patients had sUPAR levels just below the cut-off, while the group ‘c’ patients had plasma sUPAR levels higher than cut-off. (B) – (E) Heatmaps depicting changes in TPM values of selected differentially expressed genes. The genes belong to one of the four indicated groups, categorised on the basis of pattern of changes in expression in patients with increasing plasma sUPAR levels. The genes belonging to the threshold up/downregulated categories include genes which show significant up/downregulation in the group c patients as compared to the group b patients, while either showing insignificant changes or significant changes in the opposite direction when comparing group b to group a. On the other hand, genes belonging to the gradient up/downregulated categories include those which show significant gradual/stepwise up/downregulation from group a to group c, through group b. The TPM value for each gene was calculated as the average of the TPM values of all the significant differentially expressed transcripts. (F) – (I) Radar plots each depicting selected enriched pathways (for threshold/gradient up/downregulated genes) as determined from the Reactome database using the NetworkAnalyst software. The values represent the ratio of number of hits (genes) obtained in our dataset as compared to the total number of genes implicated in each pathway in the database. (J) shows the list of pathways denoted as numbers (1-20) in the radar plots.

In case of immune transcriptome also we found that a subset of genes was differentially regulated showing expression distribution in a gradient across the plasma sUPAR concentrations (Figure 7B, C, F, G). Major pathways enriched for genes with upregulated expression with increasing sUPAR concentration were signaling cascades for toll-like receptor (TLR) 4, TLR5 and IL1 receptor, pathway of cross-presentation of soluble exogenous antigens and eNOS activation pathway (Figure 7F). The pathways enriched for genes with decreasing expression across the increasing sUPAR gradient were mostly concerning type I interferon (IFN) responses to the virus, viz. pathways involving DNA-dependent activator of IFN-regulatory factors (DAI) pathway, TLR3 activation pathway, cytosolic DNA-sensing pathway as well as pathways involved in antiviral mechanisms by IFN-stimulated genes and IRF3-mediated activation of type I IFNs (Figure 7G). Thus the gene expression changes represented a deficiency in type I IFN mediated antiviral mechanisms and ramped up systemic inflammation. Figure 7B and C shows the major genes that represent these pathways.

On the other hand, another subset of differentially expressed genes showed major changes across the cut-off value, which we called threshold up- or downregulation (Figure 7D, E, H, I). Major pathways enriched by this subset of upregulated genes were involved in UPAR signaling, coagulation cascade and platelet functions (Figure 7D, H), confirming our insights gathered from the proteomics studies. It also included pathways involving T cell activation like antigen presentation by MHC class II and CD28 costimulatory pathways. Among the downregulated pathways were MHC class I antigen presentation, platelet degranulation and the signaling cascade downstream of TLR1/2 and TLR6/2 (Figure 7E, I).

### Association of plasma sUPAR with clinical outcomes in severe COVID-19

To further validate the association of plasma sUPAR level with the clinical outcomes in severe COVID-19 patients and its potential as a risk-strtifier, we compared 30 day survival and time to disease remission (getting discharged from the hospital) between the ARDS patients with plasma sUPAR levels lower than the derived cut-off value (designated sUPAR_lo_) and the ones with higher than cut-off sUPAR levels (designated sUPAR_hi_). The sUPAR_lo_ patients were found to have significant survival benefit in Kaplan Meier curve analysis as well as significantly faster remission, with median duration = 13 days for sUPAR_lo_ patients compared to 25 days for the sUPAR_hi_ group (Figure 8A, B).

**Fig. 8.**
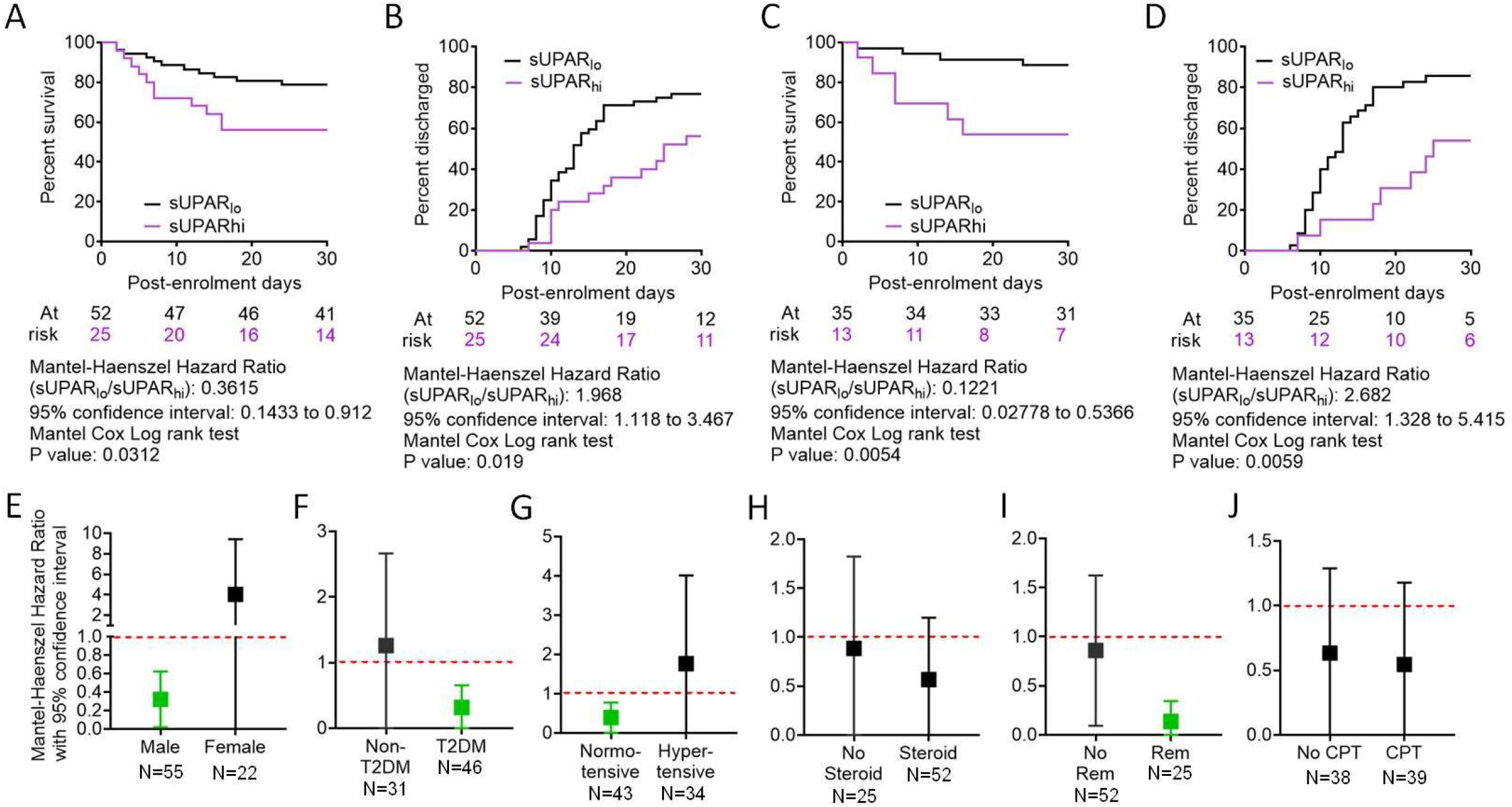
Comparison of clinical outcomes between severe COVID-19 patients with low and high plasma sUPAR levels. (A) and (C) Survival of patients in the two arms from the day of enrolment till day 30 post-enrolment are compared in a Kaplan-Meier curve, for all age groups (A) as well as for patients aged <65 years (B). Surviving patients were censored on day 30 post-enrolment. (B) and (D) Hospital stay duration of the patients from both groups (sUPAR_lo_ and sUPAR_hi_) since the day of enrolment are plotted in an ascending Kaplan-Meier curve, for all age groups (C) as well as for patients aged <65 years (D). (E) – (I) Forest plots of hazard ratio for survival benefits registered in sUPAR_lo_ patients, compared between males and females (E), patients with or without type 2 diabetes mellitus (F), patients with or without hypertension (G), treatment groups of corticosteroids (H) and remdesivir (I), across all age-groups. Deaths and non-remission at day 30 post-enrolment were censored. For all outcomes comparisons Mantel Cox log rank test was performed. For all comparisons unpaired T tests were performed.

Finally we performed an extensive subclass analysis of the patients to get insight on better applicability of the sUPAR cut-off for predicting survival in severe COVID-19 patients. First we wanted to see if age of patients can influence the prediction efficacy, because ageing has previously been shown to be a major deterrent for respiratory pathologies and worse response to therapy [18, 19]. So we explored whether there was an enhanced survival benefit registered among younger patients in the sUPAR_lo_ group. Previous studies have indicated significant differences in COVID-19 disease outcomes based on age of the patients [20, 21], perhaps due to hypo-responsiveness of an aging lung with regard to regenerative response to tissue damage. Taking cue from these studies and also from a recent study identifying different age-trajectories that represent resilience of human health in different age-groups [22], we used an age cut-off of 65 years. It was found that the survival benefit of patients aged less than 65 years with low sUPAR levels to be way more significant and they registered even faster remission (Figure 8C, D). We also performed sub-class analyses between males and females among the ARDS patients (Figure 8E), between patients who were diabetic or hypertensive and who were not (Figure 8F, G) and patients who received different therapies, viz. corticosteroids, remdesivir and convalescent plasma (Figure 8G-I). While males, diabetics, normotensive patients and patients receiving remdesivir as part of their therapies showed statistically significant survival benefits when they were sUPAR_lo_, these sub-class analyses were handicapped by lower sample sizes for the sub-classes and thus warrant further exploration in bigger cohorts of severe COVID-19 patients.

## DISCUSSION

Identification of high plasma levels of soluble UPAR in severe COVID-19 patients and association of lower plasma levels of sUPAR with favorable clinical outcomes offer a strong risk-stratifier, which will be of great translational value for effective triage and optimal timing for critical care in the patients. As discussed before, UPAR plays a regulatory role in both hemostatic pathways and complement pathway [10-13]. In our study insights gathered from proteomics and transcriptomics studies also pointed to association of a state of hyper-coagulation as well as complement activation with increasing sUPAR levels. It was evident from the systemic depletion of coagulation factors like thrombin and factor X, the deficiency of regulatory proteins that inhibit coagulation and a systemic dysbalance between factors that favour and inhibit complement activation. Decrease in plasma abundance of albumin may be indicative of an ongoing systemic or localized vascular leakage, which is known to be associated with the hypercoagulable states associated with systemic inflammations [23]. The noted upward abundance across a threshold of the haemoglobin alpha and beta chains, which are normally not expected to be abundant in plasma, points to the possibility of a concomitantly active haemolytic mechanism. The state of increased complement activation can be linked to this possibility, though it warrants further mechanistic exploration. Notably, such occurrences are already reported in patients with COVID-19 [24-26]. Moreover, sUPAR has been shown to target the FPR1 receptor, expressed dominantly in neutrophils but also on other immune cells, and thus can play a major role in chemotactic migration as well as proinflammatory cytokine induction through signaling downstream of FPR1 [10]. Interaction between UPAR and complement receptor 3 is known to regulate phagocytosis by neutrophils [13].

A previous study reporting higher sUPAR plasma levels in critically ill patients found a healthy median plasma level of 2100 pg/ml and also noted its dominant expression in myeloid cells (in this case neutrophils) [27]. Similar steady-state plasma levels were also reported by studies noting higher sUPAR levels in patients with COPD [28] and community-acquired pneumonia [29]. Thus the cut-off plasma level associated with favourable outcomes derived from the present Indian cohort of COVID-19 patients, which was 1996.809pg/ml, conforms to previous assessments in other cohorts. Of note here, a previous study with two small cohorts of severe COVID-19 patients from Greece and USA derived a cut-off of 6000pg/ml to be a strong predictor for progression to severe respiratory failure [14]. Nevertheless ethnic differences are expected to affect the steady-state of sUPAR to a great extent and further studies are warranted in different ethnicities, which may also provide insights on variable susceptibility to COVID-19 severity. Better risk stratification by plasma sUPAR in patients aged less than 65 years from our cohort further highlights the relative deficiency in terms of resolution of tissue damage in ageing lungs [18, 19].

Another important consideration for potential of sUPAR as a risk-stratifying biomarker is the long stability of sUPAR in plasma samples, which makes it a useful biomarker for operational issues as well [30]. Thus the present study identified soluble UPAR as a useful biomarker for prognostic stratification of COVID-19 patients who have progressed to ARDS in an Indian cohort. ARDS pathophysiology in COVID-19 is increasingly being appreciated in terms of alveolar inflammation-associated pulmonary intravascular coagulation [31, 32]. As UPAR-expressing myeloid cells are prevalent in both circulation and pulmonary tissue spaces, soluble UPAR may be a key link between the abnormally expanded circulating myeloid cell compartment in severe COVID-19 patients and the systemic hyper-inflammation and hypercoagulable state encountered in these patients, which warrants further mechanistic exploration.

## MATERIALS AND METHODS

### Patient characteristics

COVID-19 patients with mild symptoms (N=8) or ARDS (N=77) were recruited at the ID & BG Hospital, Kolkata, India (detailed group-wise characteristics are depicted in supplemental table 1). Peripheral blood sampling in EDTA was done on the day of enrolment with due ethical approval from the institutional review boards of ID & BG Hospital, Kolkata, India (IDBGH/Ethics/2429) and CSIR-Indian Institute of Chemical Biology, Kolkata, India (IICB/IRB/2020/3P), in accordance with the Helsinki Declaration. The ARDS patients were recruited as part of a randomised control trial (CTRI/2020/05/025209) which has been completed and published as a preprint (https://www.medrxiv.org/content/10.1101/2020.11.25.20237883v1).

### Flow cytometry

Plasma was isolated from the EDTA blood samples by centrifugation and cryostored. The whole blood cell pellets were treated with 1 ml red blood cell (RBC) lysis buffer and the RBC-depleted leukocytes were fixed with 1% paraformaldehyde before staining with the indicated fluorophore-tagged antibodies (BD Biosciences). The stained cells were acquired in a FACS ARIA III flow cytometer and data were analyzed on FlowJo™ software.

### RNA Isolation from nasopharyngeal swab samples and RT-PCR

RNA from nasopharyngeal swab samples in TRIzol was extracted using chloroform-isopropanol method. RT-PCR for SARS-CoV-2 detection was performed using the STANDARD M nCoV Real-Time Detection kit (Cat No. 11NCO10, SD Biosensor), as per manufacturer’s protocol. The kit suggested using the cut-off of Ct value 36 for the SARS-CoV-2 genes (RdRp and E gene) and the performance of the human positive control gene to declare a sample as SARS-CoV-2 positive. CY5 labeled Internal Control is used as a positive control. CT values are presented as average of the same for the two viral genes.

### SARS-CoV-2 Surrogate Virus Neutralization Assay

Neutralizing antibodies against SARS-CoV-2 in plasma samples from COVID-19 patients were detected using GeneScript SARS-CoV-2 Surrogate Virus Neutralization kit (Cat no-L00847). Assay was performed according to manufacturer’s protocol.

### Single cell RNA sequencing data analysis

scRNA sequencing data was obtained from the publicly available GEO Datasets (accession numbers – GSE163668, GSE145926 and GSE168710 [15-17]. For GSE163668, the study involved scRNA sequencing of whole blood sample of 3 patients with severe COVID-19 (GSM4995425) and 2 patients with mild/moderate COVID-19 (GSM4995426). For GSE145926, the study involved sequencing of all cells derived from the bronchoalveolar lavage fluid of 3 mild and 6 severe Covid-19 patients. For GSE168710, the study protocol depicted that isolated monocytes from the peripheral blood of 4 healthy donors were differentiated into macrophages with M-CSF treatment. Following this, the macrophages were cultured with IFN-β,IL-4, TNF-α, and IFN-γ, in combination or alone, in the presence or absence of synovial fibroblasts as indicated in the TSNE plots, before being subjected to scRNA sequencing. We analysed the sequencing data from all 3 GEO Datasets using the Seurat R package version 4.0 [30]. ‘LogNormalize’ method was used for data normalization followed by identification of the top 4000 variable features using the ‘vst’ method and ‘FindVariableFeatures’ function. Next, ‘ScaleData’ function was used for scaling the data. Principal Component Analysis was performed on the scaled data for dimensionality reduction using the ‘RunPCA’ function, followed by clustering using the ‘FindNeighbours’ and ‘FindClusters’ functions. A TSNE plot of the data was generated using the ‘RunTSNE’ function and the ‘FeaturePlot’ function was used to depict the expression of the indicated features on the TSNE plot. Finally, the target subset of interest characterised as HLA-DRA^low^ITGAX^high^ cells was selected and the expression of ‘PLAUR’ gene in these cells was visualised on the TSNE plot. The codes are available in the Supplemental file.

### Multiplex cytokine analysis

Plasma was isolated from peripheral blood of patients collected in EDTA vials. Cytokine levels (pg/ml) were measured in cryostored plasma using the Bio-Plex Pro Human Cytokine Screening Panel 48-Plex Assay (Bio-Rad, Cat No. 12007283), using manufacturer’s protocol. Data for cytokines with detectable levels in at least 70% of the ARDS patients were analyzed and has been previously analyzed in the context of the convalescent plasma RCT, as noted before.

### ELISA for soluble UPAR in plasma

Soluble UPAR levels were measured in cryostored plasma using a ELISA kit for measuring the human protein, following manufacturer’s protocol. Quantitation of the plasma concentrations were derived from the OD values at 450nm, measured on an ELISA plate reader (Biorad), using a standard supplied by the manufacturer (Invitrogen).

### Sample preparation for proteomics analysis

10 μl plasma was diluted to 100 μl with phosphate buffer and protein precipitation was done by addition of 400 μl of acetone and incubation at 25° C for 2 min, followed by centrifugation at 10000 g for 5 min. The pellets were air dried and suspended in 100 mM Tris-HCl buffer (pH 8.5). Protein estimation was performed using Bradford assay (Sigma-Aldrich, USA). For proteomics analysis, 20 μg of protein was reduced by addition of 25 mM of dithiothreitol (Sigma-Aldrich, USA) and incubated at 60 degrees Celsius for 30 min. Cysteine alkylation was performed by addition of 55 mM iodoacetamide (Sigma-Aldrich, USA) and incubated in dark for 30 min at room temperature. Samples were digested with trypsin (V511A, Promega) with an enzyme to substrate ratio of 1:10 for 16 hours at 37 degrees Celsius. Reaction was terminated with 0.1% formic acid and dried under vacuum. Peptide clean-up was done using Oasis HLB 1cc Vac cartridge (Waters).

### Mass spectrometric proteomics analysis

DIA-SWATH analysis for the samples were done on a quadrupole-TOF hybrid mass spectrometer (TripleTOF 6600, SCIEX, USA) coupled to a nano-LC system (Eksigent NanoLC-425). 4 μg of these peptides were loaded on a trap-column (ChromXP C18CL 5 μm 120 Å, Eksigent) where desalting was performed using 0.1% formic acid in water with a flow rate of 10 μl/min for 10 min. Peptides were separated on a reverse-phase C18 analytical column (ChromXP C18, 3 μm 120 Å, Eksigent) in a 57 minutes gradient of buffer A (0.1% formic acid in water) and buffer B (0.1% formic in acetonitrile) at a flow rate of 5 μl/min. Buffer B was slowly increased from 3% at 0 minute to 25% in 38 min, further increased to 32% in next 5 min and ramped to 80% buffer B in next 2 min. At 0.5 min, buffer B was increased to 90% and column was washed for 2.5 minutes, buffer B was brought to initial 3% in next 1 min and column was reconditioned for next 8 min. Method with 100 precursor isolation windows were defined based on precursor m/z frequencies using the SWATH Variable Window Calculator (SCIEX), with a minimum window of 5 m/z.

### Proteomics data analysis

Data was acquired using Analyst TF 1.7.1 Software (SCIEX). Optimized source parameters were used. Ion spray voltage was set to 5.5 kV, 25 psi for the curtain gas, 35 psi for the nebulizer gas and 250 °C as source temperature. Accumulation time was set to 250 msec for the MS scan (400–1250 m/z) and 25 msec for the MS/MS scans (100–1500 m/z). Rolling collision energies were applied for each window based on the m/z range of each SWATH and a charge 2+ ion, with a collision energy spread of 5. Total cycle time was 2.8 sec. In-house spectral-ion library file (.group) was previously generated for human blood plasma proteins by searching .wiff format files generated in DDA mode against UniProtKB human FASTA database (Swissprot and TrEMBL; 74,255 entries) using Proteinpilot™Software 5.0.1 (SCIEX). A 1% global FDR at protein level, excluding shared peptides was considered for import in SWATH 2.0 microapp of PeakView 2.2 software (SCIEX). SWATH run files were added and retention time alignment was performed using peptides from abundant proteins. The processing settings for peak extraction were: maximum of 10 peptides per protein, 5 transitions per peptide, >99% peptide confidence threshold, 1% peptide FDR. XIC extraction window was set to 10 min with 75 ppm XIC Width. All information was exported in the form of MarkerView (.mrkw) files. In MarkerView 1.2.1 (SCIEX), data normalization was performed using total area sum normalization and exported to excel. Proteomics data are submitted to the PRIDE database [33].

### Receiver operator characteristic curve

The ‘cutpointr’ package in R was used to generate the receiver operating characteristic (ROC) curve and calculate the corresponding AUC value for determining the suitability and dependability of sUPAR content in the plasma of ARDS Covid patients as an indicator of their survival. The optimum threshold sUPAR value which can be used to classify survival outcome in these patients with maximum sensitivity plus specificity was also determined.

### RNA-Seq library preparation and sequencing

The RNASeq library was prepared using Illumina TruSeq Stranded Total RNA Library Prep Kit with Ribo-Zero Gold as per TruSeq Stranded Total RNA Reference Guide. We started with 250ng of the total RNA. Briefly, the cytoplasmic and mitochondrial rRNA was depleted from the total RNA, the remaining RNA was purified, fragmented and primed for cDNA synthesis. Subsequently, double stranded cDNA (ds cDNA) was synthesised, the 3’ end of the ds cDNA was adenylated to provide an overhang for adapter ligation. The IDT for Illumina - TruSeq RNA UD Indexes was used for indexing the samples to allow multiplexing, and then finally amplified and purified to enrich the adapter ligated library. The final library was quantified using Qubit™ dsDNA High Sensitivity Assay Kit (Catalog number: Q32851) and the library size was determined using Agilent High Sensitivity DNA Kit (Catalog number: 5067-4626) on Agilent Bioanalyzer 2100 platform. For sequencing, the individual library was diluted to 4nM and libraries were pooled. The pooled library was denatured using 0.2 N NaOH, and neutralized with 200mM Tris-HCl, pH 7.0. The libraries were sequenced on a Illumina NextSeq 550 platform, using high output Kit v2.5 (300 Cycles), at a final library concentration of 1.6 pM. (NextSeq 500 and NextSeq 550 SequencingSystems: Denature and Dilute Libraries Guide; Document # 15048776 v16).

### RNA-seq data processing

Filtered fastq files were processed using Salmon v1.4.0 which provides fast and bias-aware quantification of transcript expression [34]. The mapping-based mode of Salmon was used for quantification [35]. The reference transcriptome (Ensembl GRCh38, release 103) was used for indexing and quantification of the individual genes.The quantification was performed on the full transcriptome and gene-level TPM-values(transcripts per million) were computed based on the effective length of the transcripts.The TPM values are normalised for the gene length and sequencing depth and used for further analysis of differentially expressed genes.

### RNA-seq data analysis

TPM values were analysed using the online MeV software [36]. The Limma tool [37] was used to find out the differentially expressed genes (DEGs) between the 3 different groups having 3 patients each, categorised according to the concentration of sUPAR in their plasma as described in the figure legend. The list of DEGs (transcripts) (p<=0.05) provided by Limma was divided into two groups-(1) upregulated genes (having all differentially expressed transcripts upregulated) and (2) downregulated genes (having all differentially expressed transcripts downregulated) before being entered into the online NetworkAnalyst software [38] to obtain the list of enriched pathways (p<=0.05) from the Reactome database, separately for upregulated and downregulated genes. Moreover, the genes specifically implicated in each of the pathways was also obtained.

### Correlation matrix generation and visualisation

A matrix of Spearman correlation coefficients indicating the degree and directionality of association between the concentrations of the indicated parameters in plasma of COVID-19 patients was generated using the ‘Hmisc’ package in R. For visualisation of the matrix, the ‘corrplot’ package in R was used.

### Statistics

All statistical analyses, as depicted in the results as well in appropriate figures and their legends, were performed using Graphpad Prism 8 or in some cases using R. In all cases Spearman correlation analysis and Mann Whitney test was performed unless otherwise stated. Primary outcomes of survival and disease remission (in terms of time till discharge from hospital) were compared between the two arms using Kaplan Meier Curve analysis - Mantel-Haenszel Hazard Ratio was calculated and statistical significance was tested by Mantel-Cox log rank test.

## Data Availability

The mass spectrometry proteomics data have been deposited to the ProteomeXchange Consortium via the PRIDE partner repository with the dataset identifier PXD026510. Detailed pathway enrichment data derived from the transcriptome analysis are depicted in supplemental table 4. Codes for reanalysis of the single cell RNA sequencing data are provided in supplemental information. All other data pertaining to the manuscript are available with the corresponding authors.

## Author contributions

J.S., R.D., P.B., M.A.H. and B.P.S. performed the ELISA, multiplex cytokine assay and flow cytometry; R.D., P.B. and S.C. did flow cytometric analysis; S.R.P., Y.R., K.C., S.B., D.M. B.S., B.S.S and P.B. recruited patients and participated in the clinical management of the patients; P.S., M.K. and S.S. performed the mass spectrometric proteomics; P.M., S.S, P.D., P.C. and R.P. performed the next generation sequencing and analysis; D.R.C. performed scRNAseq, RNAseq and ROC analysis; S.P. and P.G. helped with data analysis; D.G. conceptualized the study; D.G. and S.S. designed the experiments; D.G. wrote the manuscript.

## Data availability

The mass spectrometry proteomics data have been deposited to the ProteomeXchange Consortium via the PRIDE partner repository with the dataset identifier PXD026510. Detailed pathway enrichment data derived from the transcriptome analysis are depicted in supplemental table 4. Codes for reanalysis of the single cell RNA sequencing data are provided in supplemental information.

