## Supplemental for "Plasma gradient of soluble urokinase-type plasminogen activator receptor is linked to pathogenic plasma proteome and immune transcriptome and stratifies outcomes in severe COVID-19"

Jafar Sarif et al.

### Supplemental information

Supplemental table 1-4

Supplemental computing codes

Supplemental references 39-71

**Supplemental table 1:** Demography, co-morbidities and pharmacotherapy among the COVID-19 patients with ARDS

| | All ARDS | Low sUPAR | High sUPAR |
|---|---|---|---|
| **Demography** | | | |
| Male | N=55, 71.43 %, Age=62±12.12 years | N=36, 46.75%, Age=59.5± 11.98 years | N=19, 24.68%, Age=69±12.41 years |
| Female | N=22, 28.57%, Age=59.5±10.03 years | N=16, 20.78%, Age=59.5±10.22 years | N=6, 7.79%, Age=59.5±10.45 years |
| **Major co-morbidities** | | | |
| Type 2 diabetes | N=46, 59.74% | N=33, 42.86%% | N13=, 16.88% |
| Hypertension | N=34, 44.16% | N=27, 35.06% | N=7, 9.09% |
| **Pharmacotherapy** | | | |
| Standard-of-care | N=77, 100% | N=52, 100% | N=25, 100% |
| Corticosteroids | N=52, 67.53% | N=39, 50.65% | N=13, 16.88% |
| Remdesivir | N=25, 32.47% | N=20, 25.97% | N=5, 6.49% |
| Convalescent plasma | N=39, 50.65% | N=26, 33.77% | N=13, 16.88% |

**Supplemental table 2:** List of proteins detected in the proteomics study on plasma from severe COVID-19 patients

| Index | Peak Name | Group |
|---|---|---|
| 1 | sp\|P02768\|ALBU_HUMAN | Serum albumin OS=Homo sapiens OX=9606 GN=ALB PE=1 SV=2 |
| 2 | sp\|P04114\|APOB_HUMAN | Apolipoprotein B-100 OS=Homo sapiens OX=9606 GN=APOB PE=1 SV=2 |
| 3 | sp\|P01024\|CO3_HUMAN | Complement C3 OS=Homo sapiens OX=9606 GN=C3 PE=1 SV=2 |
| 4 | sp\|P01023\|A2MG_HUMAN | Alpha-2-macroglobulin OS=Homo sapiens OX=9606 GN=A2M PE=1 SV=3 |
| 5 | sp\|P02787\|TRFE_HUMAN | Serotransferrin OS=Homo sapiens OX=9606 GN=TF PE=1 SV=3 |
| 6 | sp\|P0C0L4\|CO4A_HUMAN | Complement C4-A OS=Homo sapiens OX=9606 GN=C4A PE=1 SV=2 |
| 7 | sp\|P00450\|CERU_HUMAN | Ceruloplasmin OS=Homo sapiens OX=9606 GN=CP PE=1 SV=1 |
| 8 | sp\|P01009\|A1AT_HUMAN | Alpha-1-antitrypsin OS=Homo sapiens OX=9606 GN=SERPINA1 PE=1 SV=3 |
| 9 | tr\|A0A0A0MS08\|A0A0A0MS08_HUMAN | Immunoglobulin heavy constant gamma 1 (Fragment) OS=Homo sapiens OX=9606 GN=IGHG1 PE=1 SV=1 |
| 10 | sp\|P02751\|FINC_HUMAN | Fibronectin OS=Homo sapiens OX=9606 GN=FN1 PE=1 SV=4 |
| 11 | sp\|P02671\|FIBA_HUMAN | Fibrinogen alpha chain OS=Homo sapiens OX=9606 GN=FGA PE=1 SV=2 |
| 12 | sp\|P08603\|CFAH_HUMAN | Complement factor H OS=Homo sapiens OX=9606 GN=CFH PE=1 SV=4 |
| 13 | sp\|P02675\|FIBB_HUMAN | Fibrinogen beta chain OS=Homo sapiens OX=9606 GN=FGB PE=1 SV=2 |
| 14 | sp\|P00738\|HPT_HUMAN | Haptoglobin OS=Homo sapiens OX=9606 GN=HP PE=1 SV=1 |
| 15 | tr\|B4E1Z4\|B4E1Z4_HUMAN | cDNA FLJ55673, highly similar to Complement factor B OS=Homo sapiens OX=9606 PE=1 SV=1 |
| 16 | sp\|P02679\|FIBG_HUMAN | Fibrinogen gamma chain OS=Homo sapiens OX=9606 GN=FGG PE=1 SV=3 |
| 17 | sp\|P00747\|PLMN_HUMAN | Plasminogen OS=Homo sapiens OX=9606 GN=PLG PE=1 SV=2 |
| 18 | tr\|B7ZKJ8\|B7ZKJ8_HUMAN | ITIH4 protein OS=Homo sapiens OX=9606 GN=ITIH4 PE=1 SV=1 |
| 19 | sp\|P02774\|VTDB_HUMAN | Vitamin D-binding protein OS=Homo sapiens OX=9606 GN=GC PE=1 SV=2 |
| 20 | sp\|P02647\|APOA1_HUMAN | Apolipoprotein A-I OS=Homo sapiens OX=9606 GN=APOA1 PE=1 SV=1 |
| 21 | sp\|P01031\|CO5_HUMAN | Complement C5 OS=Homo sapiens OX=9606 GN=C5 PE=1 SV=4 |
| 22 | sp\|P00734\|THRB_HUMAN | Prothrombin OS=Homo sapiens OX=9606 GN=F2 PE=1 SV=2 |
| 23 | tr\|Q5T985\|Q5T985_HUMAN | Inter-alpha-trypsin inhibitor heavy chain H2 OS=Homo sapiens OX=9606 GN=ITIH2 PE=1 SV=1 |
| 24 | sp\|P01871\|IGHM_HUMAN | Immunoglobulin heavy constant mu OS=Homo sapiens OX=9606 GN=IGHM PE=1 SV=4 |
| 25 | sp\|P02790\|HEMO_HUMAN | Hemopexin OS=Homo sapiens OX=9606 GN=HPX PE=1 SV=2 |
| 26 | sp\|P06727\|APOA4_HUMAN | Apolipoprotein A-IV OS=Homo sapiens OX=9606 GN=APOA4 PE=1 SV=3 |
| 27 | sp\|P01876\|IGHA1_HUMAN | Immunoglobulin heavy constant alpha 1 OS=Homo sapiens OX=9606 GN=IGHA1 PE=1 SV=2 |
| 28 | sp\|P04003\|C4BPA_HUMAN | C4b-binding protein alpha chain OS=Homo sapiens OX=9606 GN=C4BPA PE=1 SV=2 |
| 29 | sp\|P01008\|ANT3_HUMAN | Antithrombin-III OS=Homo sapiens OX=9606 GN=SERPINC1 PE=1 SV=1 |

| | | |
|---|---|---|
| 30 | sp\|P01011\|AACT_HUMAN | Alpha-1-antichymotrypsin OS=Homo sapiens OX=9606 GN=SERPINA3 PE=1 SV=2 |
| 31 | sp\|P68871\|HBB_HUMAN | Hemoglobin subunit beta OS=Homo sapiens OX=9606 GN=HBB PE=1 SV=2 |
| 32 | sp\|P19827\|ITIH1_HUMAN | Inter-alpha-trypsin inhibitor heavy chain H1 OS=Homo sapiens OX=9606 GN=ITIH1 PE=1 SV=3 |
| 33 | sp\|P10643\|CO7_HUMAN | Complement component C7 OS=Homo sapiens OX=9606 GN=C7 PE=1 SV=2 |
| 34 | sp\|P06396\|GELS_HUMAN | Gelsolin OS=Homo sapiens OX=9606 GN=GSN PE=1 SV=1 |
| 35 | sp\|P02765\|FETUA_HUMAN | Alpha-2-HS-glycoprotein OS=Homo sapiens OX=9606 GN=AHSG PE=1 SV=2 |
| 36 | sp\|P01042\|KNG1_HUMAN | Kininogen-1 OS=Homo sapiens OX=9606 GN=KNG1 PE=1 SV=2 |
| 37 | sp\|P04217\|A1BG_HUMAN | Alpha-1B-glycoprotein OS=Homo sapiens OX=9606 GN=A1BG PE=1 SV=4 |
| 38 | sp\|P02749\|APOH_HUMAN | Beta-2-glycoprotein 1 OS=Homo sapiens OX=9606 GN=APOH PE=1 SV=3 |
| 39 | sp\|P02649\|APOE_HUMAN | Apolipoprotein E OS=Homo sapiens OX=9606 GN=APOE PE=1 SV=1 |
| 40 | sp\|P05155\|IC1_HUMAN | Plasma protease C1 inhibitor OS=Homo sapiens OX=9606 GN=SERPING1 PE=1 SV=2 |
| 41 | sp\|P43652\|AFAM_HUMAN | Afamin OS=Homo sapiens OX=9606 GN=AFM PE=1 SV=1 |
| 42 | sp\|P13671\|CO6_HUMAN | Complement component C6 OS=Homo sapiens OX=9606 GN=C6 PE=1 SV=3 |
| 43 | sp\|P10909\|CLUS_HUMAN | Clusterin OS=Homo sapiens OX=9606 GN=CLU PE=1 SV=1 |
| 44 | tr\|A0A0B4J231\|A0A0B4J231_HUMAN | Immunoglobulin lambda-like polypeptide 5 OS=Homo sapiens OX=9606 GN=IGLL5 PE=1 SV=1 |
| 45 | sp\|P02763\|A1AG1_HUMAN | Alpha-1-acid glycoprotein 1 OS=Homo sapiens OX=9606 GN=ORM1 PE=1 SV=1 |
| 46 | tr\|V9GYM3\|V9GYM3_HUMAN | Apolipoprotein A-II OS=Homo sapiens OX=9606 GN=APOA2 PE=1 SV=1 |
| 47 | sp\|O75882\|ATRN_HUMAN | Attractin OS=Homo sapiens OX=9606 GN=ATRN PE=1 SV=2 |
| 48 | sp\|P25311\|ZA2G_HUMAN | Zinc-alpha-2-glycoprotein OS=Homo sapiens OX=9606 GN=AZGP1 PE=1 SV=2 |
| 49 | sp\|P04196\|HRG_HUMAN | Histidine-rich glycoprotein OS=Homo sapiens OX=9606 GN=HRG PE=1 SV=1 |
| 50 | sp\|P02760\|AMBP_HUMAN | Protein AMBP OS=Homo sapiens OX=9606 GN=AMBP PE=1 SV=1 |
| 51 | sp\|Q96PD5\|PGRP2_HUMAN | N-acetylmuramoyl-L-alanine amidase OS=Homo sapiens OX=9606 GN=PGLYRP2 PE=1 SV=1 |
| 52 | sp\|P01834\|IGKC_HUMAN | Immunoglobulin kappa constant OS=Homo sapiens OX=9606 GN=IGKC PE=1 SV=2 |
| 53 | sp\|P08697\|A2AP_HUMAN | Alpha-2-antiplasmin OS=Homo sapiens OX=9606 GN=SERPINF2 PE=1 SV=3 |
| 54 | sp\|P20742\|PZP_HUMAN | Pregnancy zone protein OS=Homo sapiens OX=9606 GN=PZP PE=1 SV=4 |
| 55 | tr\|A0A286YEY4\|A0A286YEY4_HUMAN | Immunoglobulin heavy constant gamma 2 (Fragment) OS=Homo sapiens OX=9606 GN=IGHG2 PE=1 SV=1 |
| 56 | tr\|B4DPQ0\|B4DPQ0_HUMAN | Complement C1r subcomponent OS=Homo sapiens OX=9606 GN=C1R PE=1 SV=1 |
| 57 | sp\|P09871\|C1S_HUMAN | Complement C1s subcomponent OS=Homo sapiens OX=9606 GN=C1S PE=1 SV=1 |
| 58 | sp\|P02748\|CO9_HUMAN | Complement component C9 OS=Homo sapiens OX=9606 GN=C9 PE=1 |

| | | |
|---|---|---|
| | | SV=2 |
| 59 | sp\|P01019\|ANGT_HUMAN | Angiotensinogen OS=Homo sapiens OX=9606 GN=AGT PE=1 SV=1 |
| 60 | sp\|P02766\|TTHY_HUMAN | Transthyretin OS=Homo sapiens OX=9606 GN=TTR PE=1 SV=1 |
| 61 | tr\|G3XAM2\|G3XAM2_HUMAN | Complement factor I OS=Homo sapiens OX=9606 GN=CFI PE=1 SV=1 |
| 62 | sp\|P02750\|A2GL_HUMAN | Leucine-rich alpha-2-glycoprotein OS=Homo sapiens OX=9606 GN=LRG1 PE=1 SV=2 |
| 63 | sp\|P60709\|ACTB_HUMAN | Actin, cytoplasmic 1 OS=Homo sapiens OX=9606 GN=ACTB PE=1 SV=1 |
| 64 | sp\|P01880\|IGHD_HUMAN | Immunoglobulin heavy constant delta OS=Homo sapiens OX=9606 GN=IGHD PE=1 SV=3 |
| 65 | sp\|O43866\|CD5L_HUMAN | CD5 antigen-like OS=Homo sapiens OX=9606 GN=CD5L PE=1 SV=1 |
| 66 | tr\|A0A087WW43\|A0A087WW43_HUMAN | Inter-alpha-trypsin inhibitor heavy chain H3 OS=Homo sapiens OX=9606 GN=ITIH3 PE=1 SV=1 |
| 67 | sp\|P27169\|PON1_HUMAN | Serum paraoxonase/arylesterase 1 OS=Homo sapiens OX=9606 GN=PON1 PE=1 SV=3 |
| 68 | sp\|P04004\|VTNC_HUMAN | Vitronectin OS=Homo sapiens OX=9606 GN=VTN PE=1 SV=1 |
| 69 | sp\|Q08380\|LG3BP_HUMAN | Galectin-3-binding protein OS=Homo sapiens OX=9606 GN=LGALS3BP PE=1 SV=1 |
| 70 | sp\|P36955\|PEDF_HUMAN | Pigment epithelium-derived factor OS=Homo sapiens OX=9606 GN=SERPINF1 PE=1 SV=4 |
| 71 | sp\|P08185\|CBG_HUMAN | Corticosteroid-binding globulin OS=Homo sapiens OX=9606 GN=SERPINA6 PE=1 SV=1 |
| 72 | tr\|H0YAC1\|H0YAC1_HUMAN | Plasma kallikrein (Fragment) OS=Homo sapiens OX=9606 GN=KLKB1 PE=1 SV=1 |
| 73 | sp\|P05543\|THBG_HUMAN | Thyroxine-binding globulin OS=Homo sapiens OX=9606 GN=SERPINA7 PE=1 SV=2 |
| 74 | tr\|A0A3B3ISJ1\|A0A3B3ISJ1_HUMAN | Vitamin K-dependent protein S OS=Homo sapiens OX=9606 GN=PROS1 PE=1 SV=1 |
| 75 | sp\|P51884\|LUM_HUMAN | Lumican OS=Homo sapiens OX=9606 GN=LUM PE=1 SV=2 |
| 76 | tr\|Q5VY30\|Q5VY30_HUMAN | Retinol-binding protein OS=Homo sapiens OX=9606 GN=RBP4 PE=1 SV=2 |
| 77 | sp\|P22792\|CPN2_HUMAN | Carboxypeptidase N subunit 2 OS=Homo sapiens OX=9606 GN=CPN2 PE=1 SV=3 |
| 78 | tr\|C9JF17\|C9JF17_HUMAN | Apolipoprotein D (Fragment) OS=Homo sapiens OX=9606 GN=APOD PE=1 SV=1 |
| 79 | sp\|P35858\|ALS_HUMAN | Insulin-like growth factor-binding protein complex acid labile subunit OS=Homo sapiens OX=9606 GN=IGFALS PE=1 SV=1 |
| 80 | sp\|P05546\|HEP2_HUMAN | Heparin cofactor 2 OS=Homo sapiens OX=9606 GN=SERPIND1 PE=1 SV=3 |
| 81 | tr\|A0A286YES1\|A0A286YES1_HUMAN | Immunoglobulin heavy constant gamma 3 (Fragment) OS=Homo sapiens OX=9606 GN=IGHG3 PE=1 SV=1 |
| 82 | sp\|P80108\|PHLD_HUMAN | Phosphatidylinositol-glycan-specific phospholipase D OS=Homo sapiens OX=9606 GN=GPLD1 PE=1 SV=3 |
| 83 | tr\|J3KRP0\|J3KRP0_HUMAN | Beta-Ala-His dipeptidase OS=Homo sapiens OX=9606 GN=CNDP1 PE=1 SV=2 |
| 84 | sp\|P00739\|HPTR_HUMAN | Haptoglobin-related protein OS=Homo sapiens OX=9606 GN=HPR PE=2 SV=2 |
| 85 | sp\|P19652\|A1AG2_HUMAN | Alpha-1-acid glycoprotein 2 OS=Homo sapiens OX=9606 GN=ORM2 PE=1 SV=2 |
| 86 | tr\|B0YIW2\|B0YIW2_HUMAN | Apolipoprotein C-III OS=Homo sapiens OX=9606 GN=APOC3 PE=1 SV=1 |
| 87 | tr\|F5H7G1\|F5H7G1_HUMAN | Complement component C8 beta chain OS=Homo sapiens OX=9606 GN=C8B PE=1 SV=1 |

| | | |
|---|---|---|
| 88 | sp\|P69905\|HBA_HUMAN | Hemoglobin subunit alpha OS=Homo sapiens OX=9606 GN=HBA1 PE=1 SV=2 |
| 89 | sp\|P07360\|CO8G_HUMAN | Complement component C8 gamma chain OS=Homo sapiens OX=9606 GN=C8G PE=1 SV=3 |
| 90 | sp\|P05160\|F13B_HUMAN | Coagulation factor XIII B chain OS=Homo sapiens OX=9606 GN=F13B PE=1 SV=3 |
| 91 | sp\|P07357\|CO8A_HUMAN | Complement component C8 alpha chain OS=Homo sapiens OX=9606 GN=C8A PE=1 SV=2 |
| 92 | sp\|P01591\|IGJ_HUMAN | Immunoglobulin J chain OS=Homo sapiens OX=9606 GN=JCHAIN PE=1 SV=4 |
| 93 | tr\|A0A286YFJ8\|A0A286YFJ8_HUMAN | Immunoglobulin heavy constant gamma 4 (Fragment) OS=Homo sapiens OX=9606 GN=IGHG4 PE=1 SV=1 |
| 94 | tr\|E9PHK0\|E9PHK0_HUMAN | Tetranectin OS=Homo sapiens OX=9606 GN=CLEC3B PE=1 SV=1 |
| 95 | sp\|P80748\|LV321_HUMAN | Immunoglobulin lambda variable 3-21 OS=Homo sapiens OX=9606 GN=IGLV3-21 PE=1 SV=2 |
| 96 | tr\|A0A096LPE2\|A0A096LPE2_HUMAN | SAA2-SAA4 readthrough OS=Homo sapiens OX=9606 GN=SAA2-SAA4 PE=4 SV=1 |
| 97 | sp\|P04264\|K2C1_HUMAN | Keratin, type II cytoskeletal 1 OS=Homo sapiens OX=9606 GN=KRT1 PE=1 SV=6 |
| 98 | tr\|K7ER74\|K7ER74_HUMAN | APOC4-APOC2 readthrough (NMD candidate) OS=Homo sapiens OX=9606 GN=APOC4-APOC2 PE=1 SV=1 |
| 99 | sp\|A0A0B4J1Y9\|HV372_HUMAN | Immunoglobulin heavy variable 3-72 OS=Homo sapiens OX=9606 GN=IGHV3-72 PE=3 SV=1 |
| 100 | sp\|P43251\|BTD_HUMAN | Biotinidase OS=Homo sapiens OX=9606 GN=BTD PE=1 SV=2 |
| 101 | tr\|A0A0J9YY99\|A0A0J9YY99_HUMAN | Uncharacterized protein (Fragment) OS=Homo sapiens OX=9606 PE=1 SV=1 |
| 102 | sp\|P29622\|KAIN_HUMAN | Kallistatin OS=Homo sapiens OX=9606 GN=SERPINA4 PE=1 SV=3 |
| 103 | sp\|O75636\|FCN3_HUMAN | Ficolin-3 OS=Homo sapiens OX=9606 GN=FCN3 PE=1 SV=2 |
| 104 | sp\|O14791\|APOL1_HUMAN | Apolipoprotein L1 OS=Homo sapiens OX=9606 GN=APOL1 PE=1 SV=5 |
| 105 | sp\|P04275\|VWF_HUMAN | von Willebrand factor OS=Homo sapiens OX=9606 GN=VWF PE=1 SV=4 |
| 106 | sp\|P01619\|KV320_HUMAN | Immunoglobulin kappa variable 3-20 OS=Homo sapiens OX=9606 GN=IGKV3-20 PE=1 SV=2 |
| 107 | sp\|P01599\|KV117_HUMAN | Immunoglobulin kappa variable 1-17 OS=Homo sapiens OX=9606 GN=IGKV1-17 PE=1 SV=2 |
| 108 | sp\|P23083\|HV102_HUMAN | Immunoglobulin heavy variable 1-2 OS=Homo sapiens OX=9606 GN=IGHV1-2 PE=1 SV=2 |
| 109 | sp\|P02747\|C1QC_HUMAN | Complement C1q subcomponent subunit C OS=Homo sapiens OX=9606 GN=C1QC PE=1 SV=3 |
| 110 | sp\|P00915\|CAH1_HUMAN | Carbonic anhydrase 1 OS=Homo sapiens OX=9606 GN=CA1 PE=1 SV=2 |
| 111 | sp\|P02042\|HBD_HUMAN | Hemoglobin subunit delta OS=Homo sapiens OX=9606 GN=HBD PE=1 SV=2 |
| 112 | sp\|P00748\|FA12_HUMAN | Coagulation factor XII OS=Homo sapiens OX=9606 GN=F12 PE=1 SV=3 |
| 113 | tr\|I3L145\|I3L145_HUMAN | Sex hormone-binding globulin OS=Homo sapiens OX=9606 GN=SHBG PE=1 SV=1 |
| 114 | sp\|P01714\|LV319_HUMAN | Immunoglobulin lambda variable 3-19 OS=Homo sapiens OX=9606 GN=IGLV3-19 PE=1 SV=2 |
| 115 | sp\|P01782\|HV309_HUMAN | Immunoglobulin heavy variable 3-9 OS=Homo sapiens OX=9606 GN=IGHV3-9 PE=1 SV=2 |
| 116 | sp\|P0DOY2\|IGLC2_HUMAN | Immunoglobulin lambda constant 2 OS=Homo sapiens OX=9606 GN=IGLC2 PE=1 SV=1 |

| | | |
|---|---|---|
| 117 | tr\|H9KV75\|H9KV75_HUMAN | Alpha-actinin-1 OS=Homo sapiens OX=9606 GN=ACTN1 PE=1 SV=1 |
| 118 | sp\|P02743\|SAMP_HUMAN | Serum amyloid P-component OS=Homo sapiens OX=9606 GN=APCS PE=1 SV=2 |
| 119 | tr\|K7ERI9\|K7ERI9_HUMAN | Apolipoprotein C-I (Fragment) OS=Homo sapiens OX=9606 GN=APOC1 PE=1 SV=1 |
| 120 | sp\|O95445\|APOM_HUMAN | Apolipoprotein M OS=Homo sapiens OX=9606 GN=APOM PE=1 SV=2 |
| 121 | sp\|P00488\|F13A_HUMAN | Coagulation factor XIII A chain OS=Homo sapiens OX=9606 GN=F13A1 PE=1 SV=4 |
| 122 | sp\|P01624\|KV315_HUMAN | Immunoglobulin kappa variable 3-15 OS=Homo sapiens OX=9606 GN=IGKV3-15 PE=1 SV=2 |
| 123 | sp\|P04180\|LCAT_HUMAN | Phosphatidylcholine-sterol acyltransferase OS=Homo sapiens OX=9606 GN=LCAT PE=1 SV=1 |
| 124 | sp\|A0A0C4DH38\|HV551_HUMAN | Immunoglobulin heavy variable 5-51 OS=Homo sapiens OX=9606 GN=IGHV5-51 PE=3 SV=1 |
| 125 | sp\|P20851\|C4BPB_HUMAN | C4b-binding protein beta chain OS=Homo sapiens OX=9606 GN=C4BPB PE=1 SV=1 |
| 126 | sp\|A0A0A0MS15\|HV349_HUMAN | Immunoglobulin heavy variable 3-49 OS=Homo sapiens OX=9606 GN=IGHV3-49 PE=3 SV=1 |
| 127 | sp\|P00742\|FA10_HUMAN | Coagulation factor X OS=Homo sapiens OX=9606 GN=F10 PE=1 SV=2 |
| 128 | tr\|D6R934\|D6R934_HUMAN | Complement C1q subcomponent subunit B OS=Homo sapiens OX=9606 GN=C1QB PE=1 SV=1 |
| 129 | sp\|Q9NZP8\|C1RL_HUMAN | Complement C1r subcomponent-like protein OS=Homo sapiens OX=9606 GN=C1RL PE=1 SV=2 |
| 130 | tr\|A0A087X0Q4\|A0A087X0Q4_HUMAN | Immunoglobulin kappa variable 2-40 OS=Homo sapiens OX=9606 GN=IGKV2-40 PE=1 SV=1 |
| 131 | sp\|P0C0L5\|CO4B_HUMAN | Complement C4-B OS=Homo sapiens OX=9606 GN=C4B PE=1 SV=2 |
| 132 | sp\|P01742\|HV169_HUMAN | Immunoglobulin heavy variable 1-69 OS=Homo sapiens OX=9606 GN=IGHV1-69 PE=1 SV=2 |
| 133 | tr\|B1AHL2\|B1AHL2_HUMAN | Fibulin-1 OS=Homo sapiens OX=9606 GN=FBLN1 PE=1 SV=1 |
| 134 | sp\|P01700\|LV147_HUMAN | Immunoglobulin lambda variable 1-47 OS=Homo sapiens OX=9606 GN=IGLV1-47 PE=1 SV=2 |
| 135 | sp\|P01602\|KV105_HUMAN | Immunoglobulin kappa variable 1-5 OS=Homo sapiens OX=9606 GN=IGKV1-5 PE=1 SV=2 |
| 136 | tr\|F8WF14\|F8WF14_HUMAN | Carboxylic ester hydrolase OS=Homo sapiens OX=9606 GN=BCHE PE=1 SV=1 |
| 137 | sp\|P06312\|KV401_HUMAN | Immunoglobulin kappa variable 4-1 OS=Homo sapiens OX=9606 GN=IGKV4-1 PE=1 SV=1 |
| 138 | tr\|B1AKG0\|B1AKG0_HUMAN | Complement factor H-related protein 1 OS=Homo sapiens OX=9606 GN=CFHR1 PE=1 SV=1 |
| 139 | tr\|A0A0A0MRJ7\|A0A0A0MRJ7_HUMAN | Coagulation factor V OS=Homo sapiens OX=9606 GN=F5 PE=1 SV=1 |
| 140 | sp\|P04433\|KV311_HUMAN | Immunoglobulin kappa variable 3-11 OS=Homo sapiens OX=9606 GN=IGKV3-11 PE=1 SV=1 |
| 141 | sp\|P01772\|HV333_HUMAN | Immunoglobulin heavy variable 3-33 OS=Homo sapiens OX=9606 GN=IGHV3-33 PE=1 SV=2 |
| 142 | tr\|E9PFZ2\|E9PFZ2_HUMAN | Ceruloplasmin OS=Homo sapiens OX=9606 GN=CP PE=1 SV=1 |
| 143 | sp\|A0A0C4DH33\|HV124_HUMAN | Immunoglobulin heavy variable 1-24 OS=Homo sapiens OX=9606 GN=IGHV1-24 PE=3 SV=1 |
| 144 | sp\|Q14520\|HABP2_HUMAN | Hyaluronan-binding protein 2 OS=Homo sapiens OX=9606 GN=HABP2 PE=1 SV=1 |
| 145 | sp\|P04432\|KVD39_HUMA | Immunoglobulin kappa variable 1D-39 OS=Homo sapiens OX=9606 |

| | | |
|---|---|---|
| | N | GN=IGKV1D-39 PE=3 SV=2 |
| 146 | sp\|P0DP07\|HV431_HUMAN | Immunoglobulin heavy variable 4-31 OS=Homo sapiens OX=9606 GN=IGHV4-31 PE=3 SV=1 |
| 147 | sp\|P35908\|K22E_HUMAN | Keratin, type II cytoskeletal 2 epidermal OS=Homo sapiens OX=9606 GN=KRT2 PE=1 SV=2 |
| 148 | sp\|P13645\|K1C10_HUMAN | Keratin, type I cytoskeletal 10 OS=Homo sapiens OX=9606 GN=KRT10 PE=1 SV=6 |
| 149 | sp\|P08519\|APOA_HUMAN | Apolipoprotein(a) OS=Homo sapiens OX=9606 GN=LPA PE=1 SV=1 |
| 150 | sp\|P01721\|LV657_HUMAN | Immunoglobulin lambda variable 6-57 OS=Homo sapiens OX=9606 GN=IGLV6-57 PE=1 SV=2 |
| 151 | sp\|P06681\|CO2_HUMAN | Complement C2 OS=Homo sapiens OX=9606 GN=C2 PE=1 SV=2 |
| 152 | sp\|P08571\|CD14_HUMAN | Monocyte differentiation antigen CD14 OS=Homo sapiens OX=9606 GN=CD14 PE=1 SV=2 |
| 153 | sp\|O95497\|VNN1_HUMAN | Pantetheinase OS=Homo sapiens OX=9606 GN=VNN1 PE=1 SV=2 |
| 154 | tr\|G3V0E5\|G3V0E5_HUMAN | Transferrin receptor (P90, CD71), isoform CRA_c OS=Homo sapiens OX=9606 GN=TFRC PE=1 SV=1 |
| 155 | tr\|Q5VVP7\|Q5VVP7_HUMAN | C-reactive protein OS=Homo sapiens OX=9606 GN=CRP PE=1 SV=1 |
| 156 | sp\|A0A0C4DH69\|KV109_HUMAN | Immunoglobulin kappa variable 1-9 OS=Homo sapiens OX=9606 GN=IGKV1-9 PE=3 SV=1 |
| 157 | tr\|A0A087WXI2\|A0A087WXI2_HUMAN | IgGFc-binding protein (Fragment) OS=Homo sapiens OX=9606 GN=FCGBP PE=1 SV=2 |
| 158 | tr\|A0A024R6I7\|A0A024R6I7_HUMAN | Alpha-1-antitrypsin OS=Homo sapiens OX=9606 GN=SERPINA1 PE=1 SV=1 |
| 159 | sp\|A0A075B6K5\|LV39_HUMAN | Immunoglobulin lambda variable 3-9 OS=Homo sapiens OX=9606 GN=IGLV3-9 PE=3 SV=1 |
| 160 | sp\|A0A0C4DH68\|KV224_HUMAN | Immunoglobulin kappa variable 2-24 OS=Homo sapiens OX=9606 GN=IGKV2-24 PE=3 SV=1 |
| 161 | sp\|A0M8Q6\|IGLC7_HUMAN | Immunoglobulin lambda constant 7 OS=Homo sapiens OX=9606 GN=IGLC7 PE=1 SV=3 |
| 162 | sp\|A0A0B4J1X5\|HV374_HUMAN | Immunoglobulin heavy variable 3-74 OS=Homo sapiens OX=9606 GN=IGHV3-74 PE=3 SV=1 |
| 163 | tr\|A0A0C4DH35\|A0A0C4DH35_HUMAN | Immunoglobulin heavy variable 3-35 (non-functional) (Fragment) OS=Homo sapiens OX=9606 GN=IGHV3-35 PE=1 SV=1 |
| 164 | tr\|A0A0G2JSC0\|A0A0G2JSC0_HUMAN | Immunoglobulin lambda variable 5-45 (Fragment) OS=Homo sapiens OX=9606 GN=IGLV5-45 PE=1 SV=1 |
| 165 | tr\|A0A0G2JMB2\|A0A0G2JMB2_HUMAN | Immunoglobulin heavy constant alpha 2 (Fragment) OS=Homo sapiens OX=9606 GN=IGHA2 PE=1 SV=1 |
| 166 | sp\|A0A075B6K4\|LV310_HUMAN | Immunoglobulin lambda variable 3-10 OS=Homo sapiens OX=9606 GN=IGLV3-10 PE=3 SV=2 |
| 167 | sp\|P01766\|HV313_HUMAN | Immunoglobulin heavy variable 3-13 OS=Homo sapiens OX=9606 GN=IGHV3-13 PE=1 SV=2 |
| 168 | sp\|P01743\|HV146_HUMAN | Immunoglobulin heavy variable 1-46 OS=Homo sapiens OX=9606 GN=IGHV1-46 PE=1 SV=2 |
| 169 | sp\|A0A0C4DH31\|HV118_HUMAN | Immunoglobulin heavy variable 1-18 OS=Homo sapiens OX=9606 GN=IGHV1-18 PE=3 SV=1 |
| 170 | tr\|A0A075B7D0\|A0A075B7D0_HUMAN | Immunoglobulin heavy variable 1/OR15-1 (non-functional) (Fragment) OS=Homo sapiens OX=9606 GN=IGHV1OR15-1 PE=1 SV=1 |
| 171 | tr\|E9PQD6\|E9PQD6_HUMAN | Serum amyloid A protein OS=Homo sapiens OX=9606 GN=SAA1 PE=1 SV=1 |
| 172 | tr\|A0A2Q2TTZ9\|A0A2Q2TTZ9_HUMAN | Immunoglobulin kappa variable 1-33 OS=Homo sapiens OX=9606 GN=IGKV1D-33 PE=1 SV=1 |
| 173 | tr\|A0A075B7B8\|A0A075B | Immunoglobulin heavy variable 3/OR16-12 (non-functional) |

| | | |
|---|---|---|
| | 7B8_HUMAN | (Fragment) OS=Homo sapiens OX=9606 GN=IGHV3OR16-12 PE=1 SV=1 |
| 174 | sp\|A0A0C4DH34\|HV428_HUMAN | Immunoglobulin heavy variable 4-28 OS=Homo sapiens OX=9606 GN=IGHV4-28 PE=3 SV=1 |
| 175 | tr\|H0YCQ7\|H0YCQ7_HUMAN | Glutamine and serine-rich protein 1 (Fragment) OS=Homo sapiens OX=9606 GN=QSER1 PE=1 SV=1 |
| 176 | tr\|E5RGB5\|E5RGB5_HUMAN | Bridging integrator 3 OS=Homo sapiens OX=9606 GN=BIN3 PE=4 SV=1 |
| 177 | tr\|C9J6K0\|C9J6K0_HUMAN | Secreted phosphoprotein 24 (Fragment) OS=Homo sapiens OX=9606 GN=SPP2 PE=1 SV=1 |
| 178 | tr\|A0A2R8YGX3\|A0A2R8YGX3_HUMAN | Tropomyosin alpha-4 chain OS=Homo sapiens OX=9606 GN=TPM4 PE=1 SV=1 |
| 179 | sp\|Q9HAK2\|COE2_HUMAN | Transcription factor COE2 OS=Homo sapiens OX=9606 GN=EBF2 PE=2 SV=4 |

**Supplemental table 3: Proteins showing significantly correlated plasma abundance with plasma sUPAR concentrations**

| Protein | Correlation with plasma sUPAR concentration | Functional attributes and potential role in pathology | Projected role in disease outcome |
|---|---|---|---|
| FIBA | Monotonic, positive | FIBA or alpha fibrinogen, on cleaved by thrombin generates fibrin, the most abundant protein in a blood clot. | Unfavorable |
| D6R934 (C1QB) | Monotonic, positive | C1QB or Complement C1Q subunit B, is a critical component of the classical complement pathway, thus has both tissue-sparing as well as tissue damaging effects depending on contexts of infections, it has been shown to upregulated in myeloid cells in COVID-19 patients [39]. | Unfavorable |
| PHLD | Threshold effect, positive | Phosphatidylinositol-glycan-specific phospholipase D hydrolyses GPI-anchors of several inflammatory proteins upon hydrolysis of proinflammatory cytokines like IL-1, TNF-α [40] | Unfavorable |
| HABP2 | Monotonic, positive | HABP2 or Hyaluronan-binding protein 2, also known as FSAP or factor VII activating protease is a critical coagulation factor [41, 42]. Increased abundance of HABP2 is known in sepsis [43]. | Unavorable |
| B7ZKJ8 (ITIH4) | Monotonic, positive | ITIH4 or Inter-alpha-trypsin inhibitor heavey chain 4, is upregulated in inflammatory contexts, mainly in response to IL-6 signaling [44]. It has also shown to upregulate in COPD [45]. Its anti-inflammatory role is evident from its protease inhibitory function and inhibition of MASP-1 and plasma kallikrein [46]. | Favorable |
| APOH | Monotonic, positive | APOH or Apolipoprotein H, also known as beta-2 glycoprotein 1, plays a role in coagulation [47, 48] with predominantly anti-coagulatory function, has been shown to increase in abundance in inflammatory contexts like COPD and SLE. | Favorable |
| K7ERI9 and K7ER74 (APOC1 and APOC4-2) | Monotonic, positive | Apolipoprotein C1 as well as C4 and C2 associates with triglyceride-rich lipoproteins and HDL and thus is critical in lipid metabolism and play a role in antiinflamatory role of HDL [49]. In addition APOC1 is known to be reduced in sepsis associated inflammation and shown to seclude bacterial TLR4 ligands in such contexts [50, 51]. | Favorable |
| E5RGB5 (BIN3) | Monotonic, positive | BIN3 or Bridging integrator 3, is a protein implicated in endocytic pathways [52]. | Favorable? |
| CAH1 | Threshold effect, | CAH1 or Carbonic anhydrase 1 is member of a | Unfavorable |

| | | | |
|---|---|---|---|
| | positive | ubiquitously expressed family of enzymes that catalyze reversible hydration of carbon dioxide to bicarbonate and protons. Interestingly a pro-inflammatory role of CAH is known in macrophages as well as in mast cells [53, 54]. | |
| RBP | Threshold effect, positive | Retinol binding protein 4 is a member of lipocalin superfamily helps in inducing proinflammatory molecule expression and also helps in lymphocyte recruitment and adherence to the endothelium by stimulating human retinal capillary endothelial cells (HRCEC) and human umbilical vein endothelial cells (HUVEC) [55]. | Unfavorable |
| AMBP | Threshold effect, positive | AMBP or Alpha-1-Microglobulin/Bikunin Precursor, an anti-inflammatory proteoglycan that stabilizes extracellular matrix by interacting with hyluronan, in combination with inter-alpha-trypsin inhibitor (ITIH1) [56, 57]. | Favorable? |
| HBA | Threshold effect, positive | Hemoglobin alpha chain, plasma hemoglobin may be indicative of a haemolytic process. | Unfavorable Hemolytic anemia? |
| HBB | Threshold effect, positive | Hemoglobin beta chain, plasma hemoglobin may be indicative of a haemolytic process. | Unfavorable Hemolytic anemia? |
| CO3 | Monotonic, negative | CO3 or Complement 3, is a key component for both classical and alternative pathways of complement activation. Proteolytic cleavage of C3 leads to the active forms which play roles in opsonization and anaphylaxis. Reduction in C3 levels has been found to be linked with worse outcomes in COVID-19 [58]. | Unfavorable |
| CO9 | Monotonic, negative | CO9 or complement component 9, is a major component of the membrane attack complex (MAC) formed as the final outcome of complement activation. Depletion of plasma CO9 may indicate its increased consumption due to recruitment of it in cell-surface MACs [59]. | Unfavorable |
| CFAH | Monotonic, negative | CFAH or complement factor H, is a regulatory glycoprotein for complement activation which prevents host cell damage by binding to C3 [60]. | Unfavorable |
| G3XAM2 (CFI) | Monotonic, negative | G3XAM2 (CFI) or Complement factor I, inhibits complement activation by cleaving C3b and C4b [61]. Genetic abnormalities of CFI have been associated with complement mediated hemolysis [62]. | Unfavorable |
| THRB | Monotonic, negative | THRB or Thrombin, is a serine protease which drives fibrinogen to fibrin conversion thus being critical for coagulation. Depletion of circulating thrombin may indicate systemic | Unfavorable |

| | | | |
|---|---|---|---|
| | | coagulopathy associated with COVID-19 [63]. | |
| FA10 | Monotonic, negative | FA10 or Factor X is the critical regulator of coagulation cascade which cleaves prothrombin to generate thrombin. Depletion of circulating FA10 may result from systemic coagulopathy associated with COVID-19. | Unfavorable |
| VTNC | Monotonic, negative | VTNC or Vitronectin is a circulating glycoprotein playing critical roles in both inhibition of coagulopathy, through inhibition of plasmaminogen activator inhibitor (PAI) as well as by protecting against complment-mediated cellular damage [64, 65]. | Unfavorable |
| ACTN1 | Monotonic, negative | ACTN1 or alpha actinin 1 is a cytoskeletal protein associated with cellular junctions with critical physiological effects in smooth muscle cells as well as epithelia and endothelial cells. Plasma level of ACTN1 may indicate smooth muscle cell injury including injury to myocardium resulting from systemic inflammation. | Favorable? |
| A1AG1 and A1AG2 | Threshold effect, negative | Alpha 1 acid glycoprotein 1 and 2, are acute phase reactants produced by liver and peripheral tissues in response to systemic inflammation and has been shown to regulate inflammatory cells and mediators with an anti-inflammatory and anti-fibrogenic role [66, 67]. | Unfavorable? |
| PLMN | Threshold effect, negative | PLMN or plasminogen is the precursor for the fibrinolytic protein plasmin. Deficiency of peptides constituent of plasminogen may point to deficiency of the fibrinolytic process leading to systemic coagulopathy, established to be associated with COVID-19 [68]. | Unfavorable |
| ANT3 | Threshold effect, negative | ANT3 or antithrombin III, is a glycoprotein with regulatory role on intrinsic pathway of coagulation mediated by inhibition of the proteases taking part in the coagulation cascade [69]. | Unfavorable |
| ALBU | Threshold effect, negative | ALBU or Albumin, depletion from plasma is indicative of vascular leakage during systemic inflammation. It has been shown to be associated with hypercoagulable state associated with ANT3 reduction encountered in systemic inflammation [70]. | Unfavorable |
| SAMP | Threshold effect, negative | SAMP or Serum amyloid P is an acute phase reactant of the pantraxin family known to have anti-inflammatory as well as anti-fibrogenic roles [71]. | Unfavorable |
| A0A096LPE2 | Threshold effect, negative | SAA2-SAA4 readthrough, serum amyloid A proteins, are acute phase reactants induced in response to systemic inflammation, which | Unfavorable |

| | | | |
|---|---|---|---|
| | | help in immune cell recruitment. | |
| APOC3 | Threshold effect, negative | APOC3 or apolipoprotein C3 is associated with VLDL particles and an increased level portends hypertriglyceridemia. | Favorable |

**Supplemetal table 4. Pathways enriched by differentially expressed genes between group ‘a’, ‘b’ and ‘c’ patients.**

**A. Enriched pathways for gradient upregulated genes**

| Pathway | Total | Expected | Hits | P.Value | FDR |
|---|---|---|---|---|---|
| Destabilization of mRNA by AUF1 (hnRNP D0) | 54 | 0.259 | 3 | 0.0021 | 0.813 |
| The citric acid (TCA) cycle and respiratory electron transport | 145 | 0.695 | 4 | 0.00478 | 0.813 |
| Deadenylation of mRNA | 24 | 0.115 | 2 | 0.00575 | 0.813 |
| Smooth Muscle Contraction | 25 | 0.12 | 2 | 0.00623 | 0.813 |
| Respiratory electron transport | 82 | 0.393 | 3 | 0.00685 | 0.813 |
| p75 NTR receptor-mediated signalling | 85 | 0.407 | 3 | 0.00757 | 0.813 |
| Regulation of mRNA Stability by Proteins that Bind AU-rich Elements | 88 | 0.422 | 3 | 0.00833 | 0.813 |
| Interactions of Tat with host cellular proteins | 2 | 0.00959 | 1 | 0.00956 | 0.813 |
| Synthesis of DNA | 95 | 0.455 | 3 | 0.0103 | 0.813 |
| Activated TLR4 signalling | 100 | 0.479 | 3 | 0.0118 | 0.813 |
| Respiratory electron transport, ATP synthesis by chemiosmotic coupling, and heat production by uncoupling proteins. | 101 | 0.484 | 3 | 0.0121 | 0.813 |
| DNA Replication | 102 | 0.489 | 3 | 0.0125 | 0.813 |
| Toll Like Receptor 4 (TLR4) Cascade | 103 | 0.494 | 3 | 0.0128 | 0.813 |
| Metabolism of mRNA | 317 | 1.52 | 5 | 0.0164 | 0.813 |
| HIV Infection | 214 | 1.03 | 4 | 0.0183 | 0.813 |
| Axonal growth stimulation | 4 | 0.0192 | 1 | 0.019 | 0.813 |
| Activation of CaMK IV | 4 | 0.0192 | 1 | 0.019 | 0.813 |
| Interleukin-1 signaling | 45 | 0.216 | 2 | 0.0194 | 0.813 |
| S Phase | 122 | 0.585 | 3 | 0.0201 | 0.813 |
| Toll-Like Receptors Cascades | 123 | 0.589 | 3 | 0.0205 | 0.813 |
| Rho GTPase cycle | 123 | 0.589 | 3 | 0.0205 | 0.813 |
| Signaling by Rho GTPases | 123 | 0.589 | 3 | 0.0205 | 0.813 |
| Metabolism of RNA | 339 | 1.62 | 5 | 0.0213 | 0.813 |
| Regulation of activated PAK-2p34 by proteasome mediated degradation | 48 | 0.23 | 2 | 0.0219 | 0.813 |
| Cross-presentation of soluble exogenous antigens | 48 | 0.23 | 2 | 0.0219 | 0.813 |

| | | | | | |
|---|---|---|---|---|---|
| (endosomes) | | | | | |
| Ubiquitin-dependent degradation of Cyclin D1 | 49 | 0.235 | 2 | 0.0227 | 0.813 |
| CDK-mediated phosphorylation and removal of Cdc6 | 49 | 0.235 | 2 | 0.0227 | 0.813 |
| Ubiquitin-dependent degradation of Cyclin D | 49 | 0.235 | 2 | 0.0227 | 0.813 |
| Regulation of ornithine decarboxylase (ODC) | 49 | 0.235 | 2 | 0.0227 | 0.813 |
| Vpu mediated degradation of CD4 | 50 | 0.24 | 2 | 0.0236 | 0.813 |
| CaMK IV-mediated phosphorylation of CREB | 5 | 0.024 | 1 | 0.0237 | 0.813 |
| Cell Cycle Checkpoints | 131 | 0.628 | 3 | 0.0242 | 0.813 |
| Deadenylation-dependent mRNA decay | 51 | 0.244 | 2 | 0.0245 | 0.813 |
| Ubiquitin Mediated Degradation of Phosphorylated Cdc25A | 52 | 0.249 | 2 | 0.0254 | 0.813 |
| p53-Independent DNA Damage Response | 52 | 0.249 | 2 | 0.0254 | 0.813 |
| p53-Independent G1/S DNA damage checkpoint | 52 | 0.249 | 2 | 0.0254 | 0.813 |
| Muscle contraction | 52 | 0.249 | 2 | 0.0254 | 0.813 |
| SCF-beta-TrCP mediated degradation of Emi1 | 53 | 0.254 | 2 | 0.0263 | 0.813 |
| Autodegradation of the E3 ubiquitin ligase COP1 | 53 | 0.254 | 2 | 0.0263 | 0.813 |
| Stabilization of p53 | 54 | 0.259 | 2 | 0.0273 | 0.813 |
| Vif-mediated degradation of APOBEC3G | 55 | 0.264 | 2 | 0.0282 | 0.813 |
| Processing of DNA ends prior to end rejoining | 6 | 0.0288 | 1 | 0.0284 | 0.813 |
| CREB phosphorylation through the activation of CaMKK | 6 | 0.0288 | 1 | 0.0284 | 0.813 |
| Host Interactions of HIV factors | 141 | 0.676 | 3 | 0.0293 | 0.813 |
| CDT1 association with the CDC6:ORC:origin complex | 57 | 0.273 | 2 | 0.0302 | 0.813 |
| SCF(Skp2)-mediated degradation of p27/p21 | 58 | 0.278 | 2 | 0.0311 | 0.813 |
| Cell Cycle | 508 | 2.43 | 6 | 0.0312 | 0.813 |
| p53-Dependent G1/S DNA damage checkpoint | 59 | 0.283 | 2 | 0.0321 | 0.813 |
| p53-Dependent G1 DNA Damage Response | 59 | 0.283 | 2 | 0.0321 | 0.813 |
| Regulation of Apoptosis | 59 | 0.283 | 2 | 0.0321 | 0.813 |
| G1/S DNA Damage Checkpoints | 62 | 0.297 | 2 | 0.0352 | 0.813 |
| Assembly of the pre-replicative complex | 63 | 0.302 | 2 | 0.0362 | 0.813 |
| ER-Phagosome pathway | 63 | 0.302 | 2 | 0.0362 | 0.813 |
| Nonhomologous End-joining (NHEJ) | 8 | 0.0383 | 1 | 0.0377 | 0.813 |
| Regulation of Insulin Secretion by Free Fatty Acids | 8 | 0.0383 | 1 | 0.0377 | 0.813 |

| | | | | | |
|---|---|---|---|---|---|
| Regulation of Insulin Secretion by Fatty Acids Bound to GPR40 (FFAR1) | 8 | 0.0383 | 1 | 0.0377 | 0.813 |
| Multifunctional anion exchangers | 8 | 0.0383 | 1 | 0.0377 | 0.813 |
| Cyclin E associated events during G1/S transition | 65 | 0.312 | 2 | 0.0384 | 0.813 |
| Degradation of beta-catenin by the destruction complex | 65 | 0.312 | 2 | 0.0384 | 0.813 |
| Signaling by Wnt | 65 | 0.312 | 2 | 0.0384 | 0.813 |
| Activation of NF-kappaB in B Cells | 66 | 0.316 | 2 | 0.0395 | 0.813 |
| Cyclin A:Cdk2-associated events at S phase entry | 66 | 0.316 | 2 | 0.0395 | 0.813 |
| Autodegradation of Cdh1 by Cdh1:APC/C | 68 | 0.326 | 2 | 0.0417 | 0.813 |
| 2-LTR circle formation | 9 | 0.0431 | 1 | 0.0423 | 0.813 |
| Axonal growth inhibition (RHOA activation) | 9 | 0.0431 | 1 | 0.0423 | 0.813 |
| eNOS activation | 9 | 0.0431 | 1 | 0.0423 | 0.813 |
| Switching of origins to a post-replicative state | 69 | 0.331 | 2 | 0.0428 | 0.813 |
| Orc1 removal from chromatin | 69 | 0.331 | 2 | 0.0428 | 0.813 |
| Cell Cycle, Mitotic | 411 | 1.97 | 5 | 0.0439 | 0.813 |
| Removal of licensing factors from origins | 71 | 0.34 | 2 | 0.045 | 0.813 |
| Regulation of DNA replication | 71 | 0.34 | 2 | 0.045 | 0.813 |
| APC/C:Cdc20 mediated degradation of Securin | 71 | 0.34 | 2 | 0.045 | 0.813 |
| Cytokine Signaling in Immune system | 286 | 1.37 | 4 | 0.0462 | 0.813 |
| Telomere Maintenance | 72 | 0.345 | 2 | 0.0462 | 0.813 |
| Methylation | 10 | 0.0479 | 1 | 0.0469 | 0.813 |
| p75NTR regulates axonogenesis | 10 | 0.0479 | 1 | 0.0469 | 0.813 |
| Signalling by NGF | 290 | 1.39 | 4 | 0.0482 | 0.813 |
| Toll Like Receptor 10 (TLR10) Cascade | 74 | 0.355 | 2 | 0.0485 | 0.813 |
| Toll Like Receptor 5 (TLR5) Cascade | 74 | 0.355 | 2 | 0.0485 | 0.813 |
| MyD88 cascade initiated on plasma membrane | 74 | 0.355 | 2 | 0.0485 | 0.813 |

**B. Enriched pathways for gradient downregulated genes**

| Pathway | Total | Expected | Hits | P.Value | FDR |
|---|---|---|---|---|---|
| NF-kB activation through FADD/RIP-1 pathway mediated by caspase-8 and -10 | 12 | 0.101 | 4 | 2.09E-06 | 0.00293 |
| DAI mediated induction of type I IFNs | 13 | 0.109 | 3 | 0.000151 | 0.0788 |

| | | | | | |
|---|---|---|---|---|---|
| TRAF3-dependent IRF activation pathway | 14 | 0.117 | 3 | 0.000191 | 0.0788 |
| RIG-I/MDA5 mediated induction of IFN-alpha/beta pathways | 67 | 0.562 | 5 | 0.000225 | 0.0788 |
| TRAF6 mediated NF-kB activation | 16 | 0.134 | 3 | 0.00029 | 0.0812 |
| Cytosolic sensors of pathogen-associated DNA | 19 | 0.159 | 3 | 0.000492 | 0.115 |
| TRAF6 mediated IRF7 activation | 28 | 0.235 | 3 | 0.00158 | 0.316 |
| Negative regulators of RIG-I/MDA5 signaling | 33 | 0.277 | 3 | 0.00255 | 0.447 |
| RIP-mediated NFkB activation via DAI | 11 | 0.0923 | 2 | 0.00362 | 0.564 |
| Activation of Chaperone Genes by XBP1(S) | 46 | 0.386 | 3 | 0.00657 | 0.916 |
| Developmental Biology | 417 | 3.5 | 9 | 0.00729 | 0.916 |
| Activation of Chaperones by IRE1alpha | 49 | 0.411 | 3 | 0.00784 | 0.916 |
| Signal regulatory protein (SIRP) family interactions | 20 | 0.168 | 2 | 0.0119 | 1 |
| Signaling by FGFR1 fusion mutants | 20 | 0.168 | 2 | 0.0119 | 1 |
| cGMP effects | 21 | 0.176 | 2 | 0.0131 | 1 |
| L1CAM interactions | 112 | 0.939 | 4 | 0.0141 | 1 |
| Export of Viral Ribonucleoproteins from Nucleus | 2 | 0.0168 | 1 | 0.0167 | 1 |
| NEP/NS2 Interacts with the Cellular Export Machinery | 2 | 0.0168 | 1 | 0.0167 | 1 |
| Unfolded Protein Response | 66 | 0.554 | 3 | 0.0176 | 1 |
| IKK complex recruitment mediated by RIP1 | 25 | 0.21 | 2 | 0.0183 | 1 |
| Antiviral mechanism by IFN-stimulated genes | 69 | 0.579 | 3 | 0.0198 | 1 |
| ISG15 antiviral mechanism | 69 | 0.579 | 3 | 0.0198 | 1 |
| Nitric oxide stimulates guanylate cyclase | 27 | 0.226 | 2 | 0.0212 | 1 |
| IRF3 mediated activation of type 1 IFN | 3 | 0.0252 | 1 | 0.025 | 1 |
| Beta oxidation of palmitoyl-CoA to myristoyl-CoA | 3 | 0.0252 | 1 | 0.025 | 1 |
| Cam-PDE 1 activation | 3 | 0.0252 | 1 | 0.025 | 1 |
| TGF-beta receptor signaling activates SMADs | 30 | 0.252 | 2 | 0.0259 | 1 |
| Signaling by FGFR1 mutants | 31 | 0.26 | 2 | 0.0275 | 1 |
| Cytokine Signaling in Immune system | 286 | 2.4 | 6 | 0.0316 | 1 |
| Plasmalogen biosynthesis | 4 | 0.0335 | 1 | 0.0331 | 1 |
| NGF processing | 4 | 0.0335 | 1 | 0.0331 | 1 |
| Neurophilin interactions with VEGF and VEGFR | 4 | 0.0335 | 1 | 0.0331 | 1 |

| | | | | | |
|---|---|---|---|---|---|
| Axon guidance | 292 | 2.45 | 6 | 0.0345 | 1 |
| TRIF-mediated TLR3/TLR4 signaling | 87 | 0.73 | 3 | 0.0361 | 1 |
| MyD88-independent cascade | 88 | 0.738 | 3 | 0.0372 | 1 |
| Toll Like Receptor 3 (TLR3) Cascade | 88 | 0.738 | 3 | 0.0372 | 1 |
| S6K1 signalling | 5 | 0.0419 | 1 | 0.0412 | 1 |
| NOSTRIN mediated eNOS trafficking | 5 | 0.0419 | 1 | 0.0412 | 1 |
| Immune System | 1140 | 9.54 | 15 | 0.0441 | 1 |
| ATM mediated phosphorylation of repair proteins | 6 | 0.0503 | 1 | 0.0493 | 1 |
| ATM mediated response to DNA double-strand break | 6 | 0.0503 | 1 | 0.0493 | 1 |
| ER Quality Control Compartment (ERQC) | 6 | 0.0503 | 1 | 0.0493 | 1 |

#### C. Enriched pathways for threshold upregulated genes

| **Pathway** | **Total** | **Expected** | **Hits** | **P.Value** | **FDR** |
|---|---|---|---|---|---|
| Attachment of GPI anchor to uPAR | 7 | 0.609 | 4 | 0.0016 | 0.612 |
| Activation of BAD and translocation to mitochondria | 17 | 1.48 | 6 | 0.00227 | 0.612 |
| Pre-NOTCH Transcription and Translation | 12 | 1.04 | 5 | 0.00231 | 0.612 |
| Bile salt and organic anion SLC transporters | 12 | 1.04 | 5 | 0.00231 | 0.612 |
| AKT-mediated inactivation of FOXO1A | 4 | 0.348 | 3 | 0.00245 | 0.612 |
| Transport of glucose and other sugars, bile salts and organic acids, metal ions and amine compounds | 95 | 8.27 | 17 | 0.00307 | 0.612 |
| L1CAM interactions | 112 | 9.75 | 19 | 0.00338 | 0.612 |
| AKT phosphorylates targets in the cytosol | 13 | 1.13 | 5 | 0.00349 | 0.612 |
| Regulation of Gene Expression by Hypoxia-inducible Factor | 9 | 0.783 | 4 | 0.00501 | 0.731 |
| Eicosanoids | 5 | 0.435 | 3 | 0.00574 | 0.731 |
| Acyl chain remodeling of DAG and TAG | 5 | 0.435 | 3 | 0.00574 | 0.731 |
| Proton-coupled neutral amino acid transporters | 2 | 0.174 | 2 | 0.00756 | 0.731 |
| Signal transduction by L1 | 35 | 3.05 | 8 | 0.00893 | 0.731 |
| Pre-NOTCH Expression and Processing | 22 | 1.91 | 6 | 0.00941 | 0.731 |
| Transmembrane transport of small molecules | 504 | 43.9 | 59 | 0.00997 | 0.731 |
| Activation of PKB | 6 | 0.522 | 3 | 0.0107 | 0.731 |
| RSK activation | 6 | 0.522 | 3 | 0.0107 | 0.731 |

| | | | | | |
|---|---|---|---|---|---|
| Negative regulation of the PI3K/AKT network | 11 | 0.957 | 4 | 0.0114 | 0.731 |
| Signaling by EGFR | 179 | 15.6 | 25 | 0.0117 | 0.731 |
| Rap1 signalling | 17 | 1.48 | 5 | 0.0125 | 0.731 |
| Intrinsic Pathway for Apoptosis | 37 | 3.22 | 8 | 0.0126 | 0.731 |
| CD28 co-stimulation | 30 | 2.61 | 7 | 0.0126 | 0.731 |
| Hemostasis | 511 | 44.5 | 59 | 0.0131 | 0.731 |
| Signaling by EGFR in Cancer | 181 | 15.7 | 25 | 0.0134 | 0.731 |
| Downstream signal transduction | 163 | 14.2 | 23 | 0.0135 | 0.731 |
| Activation of BH3-only proteins | 24 | 2.09 | 6 | 0.0146 | 0.731 |
| CTLA4 inhibitory signaling | 24 | 2.09 | 6 | 0.0146 | 0.731 |
| The role of Nef in HIV-1 replication and disease pathogenesis | 31 | 2.7 | 7 | 0.0151 | 0.731 |
| Adherens junctions interactions | 31 | 2.7 | 7 | 0.0151 | 0.731 |
| Fcgamma receptor (FCGR) dependent phagocytosis | 86 | 7.48 | 14 | 0.0157 | 0.733 |
| SLC-mediated transmembrane transport | 251 | 21.8 | 32 | 0.0173 | 0.74 |
| NrCAM interactions | 7 | 0.609 | 3 | 0.0176 | 0.74 |
| Post-transcriptional Silencing By Small RNAs | 7 | 0.609 | 3 | 0.0176 | 0.74 |
| Signaling by FGFR in disease | 178 | 15.5 | 24 | 0.0199 | 0.74 |
| CD28 dependent PI3K/Akt signaling | 19 | 1.65 | 5 | 0.0204 | 0.74 |
| Transcriptional Regulation of White Adipocyte Differentiation | 56 | 4.87 | 10 | 0.0211 | 0.74 |
| Formation of apoptosome | 3 | 0.261 | 2 | 0.0214 | 0.74 |
| Downregulation of ERBB2:ERBB3 signaling | 13 | 1.13 | 4 | 0.0215 | 0.74 |
| alpha-linolenic acid (ALA) metabolism | 13 | 1.13 | 4 | 0.0215 | 0.74 |
| alpha-linolenic (omega3) and linoleic (omega6) acid metabolism | 13 | 1.13 | 4 | 0.0215 | 0.74 |
| Kinesins | 41 | 3.57 | 8 | 0.0229 | 0.74 |
| Metabolism of lipids and lipoproteins | 507 | 44.1 | 57 | 0.024 | 0.74 |
| G beta:gamma signalling through PI3Kgamma | 27 | 2.35 | 6 | 0.0258 | 0.74 |
| Lysosome Vesicle Biogenesis | 27 | 2.35 | 6 | 0.0258 | 0.74 |
| Lysine catabolism | 8 | 0.696 | 3 | 0.0263 | 0.74 |
| Vitamin C (ascorbate) metabolism | 8 | 0.696 | 3 | 0.0263 | 0.74 |
| Nicotinate metabolism | 8 | 0.696 | 3 | 0.0263 | 0.74 |

| | | | | | |
|---|---|---|---|---|---|
| Linoleic acid (LA) metabolism | 8 | 0.696 | 3 | 0.0263 | 0.74 |
| Signaling by ERBB2 | 164 | 14.3 | 22 | 0.0264 | 0.74 |
| DAP12 signaling | 164 | 14.3 | 22 | 0.0264 | 0.74 |
| Regulation of Lipid Metabolism by Peroxisome proliferator-activated receptor alpha (PPARalpha) | 76 | 6.61 | 12 | 0.0301 | 0.826 |
| HIV Infection | 214 | 18.6 | 27 | 0.0308 | 0.826 |
| Cell-cell junction organization | 60 | 5.22 | 10 | 0.0327 | 0.826 |
| Downstream signaling of activated FGFR | 150 | 13.1 | 20 | 0.0351 | 0.826 |
| NGF signalling via TRKA from the plasma membrane | 207 | 18 | 26 | 0.0352 | 0.826 |
| Synthesis of Prostaglandins (PG) and Thromboxanes (TX) | 15 | 1.31 | 4 | 0.0357 | 0.826 |
| Signalling to p38 via RIT and RIN | 15 | 1.31 | 4 | 0.0357 | 0.826 |
| Developmental Biology | 417 | 36.3 | 47 | 0.0369 | 0.826 |
| AKT phosphorylates targets in the nucleus | 9 | 0.783 | 3 | 0.037 | 0.826 |
| Signaling by PDGF | 189 | 16.4 | 24 | 0.0376 | 0.826 |
| Signalling to ERKs | 37 | 3.22 | 7 | 0.0377 | 0.826 |
| ABC-family proteins mediated transport | 37 | 3.22 | 7 | 0.0377 | 0.826 |
| Arachidonic acid metabolism | 45 | 3.92 | 8 | 0.0381 | 0.826 |
| Signaling by SCF-KIT | 142 | 12.4 | 19 | 0.0381 | 0.826 |
| Proton-coupled monocarboxylate transport | 4 | 0.348 | 2 | 0.0403 | 0.826 |
| Signaling by FGFR | 162 | 14.1 | 21 | 0.041 | 0.826 |
| G-protein beta:gamma signalling | 30 | 2.61 | 6 | 0.0414 | 0.826 |
| Platelet activation, signaling and aggregation | 220 | 19.1 | 27 | 0.0416 | 0.826 |
| Cell junction organization | 89 | 7.74 | 13 | 0.0432 | 0.826 |
| ARMS-mediated activation | 16 | 1.39 | 4 | 0.0444 | 0.826 |
| Late Phase of HIV Life Cycle | 108 | 9.4 | 15 | 0.0463 | 0.826 |
| Transcription-coupled NER (TC-NER) | 47 | 4.09 | 8 | 0.0477 | 0.826 |
| Dual incision reaction in TC-NER | 31 | 2.7 | 6 | 0.0478 | 0.826 |
| Formation of transcription-coupled NER (TC-NER) repair complex | 31 | 2.7 | 6 | 0.0478 | 0.826 |
| Amino acid transport across the plasma membrane | 31 | 2.7 | 6 | 0.0478 | 0.826 |
| Trafficking of AMPA receptors | 31 | 2.7 | 6 | 0.0478 | 0.826 |
| Glutamate Binding, Activation of AMPA Receptors | 31 | 2.7 | 6 | 0.0478 | 0.826 |

| | | | | | |
|---|---|---|---|---|---|
| and Synaptic Plasticity | | | | | |
| Adaptive Immune System | 654 | 56.9 | 69 | 0.0478 | 0.826 |
| trans-Golgi Network Vesicle Budding | 64 | 5.57 | 10 | 0.0483 | 0.826 |
| Clathrin derived vesicle budding | 64 | 5.57 | 10 | 0.0483 | 0.826 |
| MHC class II antigen presentation | 118 | 10.3 | 16 | 0.0488 | 0.826 |
| RAF/MAP kinase cascade | 10 | 0.87 | 3 | 0.0495 | 0.826 |
| Zinc influx into cells by the SLC39 gene family | 10 | 0.87 | 3 | 0.0495 | 0.826 |
| GP1b-IX-V activation signalling | 10 | 0.87 | 3 | 0.0495 | 0.826 |

#### D. Enriched pathways for threshold downregulated genes

| Pathway | Total | Expected | Hits | P.Value | FDR |
|---|---|---|---|---|---|
| Cell Cycle | 508 | 49.4 | 71 | 0.000831 | 0.35 |
| Class I MHC mediated antigen processing & presentation | 267 | 26 | 42 | 0.00105 | 0.35 |
| Gene Expression | 1090 | 106 | 134 | 0.00115 | 0.35 |
| Nuclear Envelope Reassembly | 14 | 1.36 | 6 | 0.00125 | 0.35 |
| Initiation of Nuclear Envelope Reformation | 14 | 1.36 | 6 | 0.00125 | 0.35 |
| Cell Cycle Checkpoints | 131 | 12.7 | 24 | 0.00161 | 0.377 |
| Platelet activation, signaling and aggregation | 220 | 21.4 | 35 | 0.0022 | 0.441 |
| Immune System | 1140 | 111 | 136 | 0.00371 | 0.563 |
| Metabolism of nucleotides | 81 | 7.87 | 16 | 0.00433 | 0.563 |
| Cell Cycle, Mitotic | 411 | 39.9 | 56 | 0.00512 | 0.563 |
| Phospholipid metabolism | 135 | 13.1 | 23 | 0.00517 | 0.563 |
| N-glycan trimming in the ER and Calnexin/Calreticulin cycle | 13 | 1.26 | 5 | 0.00566 | 0.563 |
| G1/S DNA Damage Checkpoints | 62 | 6.03 | 13 | 0.00576 | 0.563 |
| Pyrimidine metabolism | 24 | 2.33 | 7 | 0.00629 | 0.563 |
| Rho GTPase cycle | 123 | 12 | 21 | 0.00718 | 0.563 |
| Signaling by Rho GTPases | 123 | 12 | 21 | 0.00718 | 0.563 |
| Metabolism of RNA | 339 | 33 | 47 | 0.00724 | 0.563 |
| Glycerophospholipid biosynthesis | 86 | 8.36 | 16 | 0.0079 | 0.563 |
| Antigen Presentation: Folding, assembly and peptide loading of class I MHC | 25 | 2.43 | 7 | 0.00802 | 0.563 |

| | | | | | |
|---|---|---|---|---|---|
| Response to elevated platelet cytosolic Ca2+ | 94 | 9.14 | 17 | 0.00838 | 0.563 |
| Deadenylation-dependent mRNA decay | 51 | 4.96 | 11 | 0.00861 | 0.563 |
| Antigen processing: Ubiquitination & Proteasome degradation | 224 | 21.8 | 33 | 0.00957 | 0.563 |
| Signaling by FGFR1 fusion mutants | 20 | 1.94 | 6 | 0.00972 | 0.563 |
| p53-Dependent G1/S DNA damage checkpoint | 59 | 5.73 | 12 | 0.01 | 0.563 |
| p53-Dependent G1 DNA Damage Response | 59 | 5.73 | 12 | 0.01 | 0.563 |
| Platelet degranulation | 89 | 8.65 | 16 | 0.011 | 0.592 |
| Clearance of Nuclear Envelope Membranes from Chromatin | 10 | 0.972 | 4 | 0.0115 | 0.598 |
| Transcription-coupled NER (TC-NER) | 47 | 4.57 | 10 | 0.0132 | 0.635 |
| Metabolism of mRNA | 317 | 30.8 | 43 | 0.0144 | 0.635 |
| Toll-Like Receptors Cascades | 123 | 12 | 20 | 0.0145 | 0.635 |
| Mitotic M-M/G1 phases | 266 | 25.9 | 37 | 0.0154 | 0.635 |
| Mitotic Anaphase | 198 | 19.2 | 29 | 0.0157 | 0.635 |
| M Phase | 233 | 22.6 | 33 | 0.0167 | 0.635 |
| ER-Phagosome pathway | 63 | 6.12 | 12 | 0.0167 | 0.635 |
| Vitamin B5 (pantothenate) metabolism | 11 | 1.07 | 4 | 0.0167 | 0.635 |
| Calnexin/calreticulin cycle | 11 | 1.07 | 4 | 0.0167 | 0.635 |
| Mitotic Metaphase and Anaphase | 199 | 19.3 | 29 | 0.0168 | 0.635 |
| Nuclear Envelope Breakdown | 17 | 1.65 | 5 | 0.0196 | 0.703 |
| G alpha (12/13) signalling events | 80 | 7.78 | 14 | 0.0209 | 0.703 |
| Regulation of mRNA Stability by Proteins that Bind AU-rich Elements | 88 | 8.55 | 15 | 0.0214 | 0.703 |
| G1/S Transition | 113 | 11 | 18 | 0.0239 | 0.703 |
| Deadenylation of mRNA | 24 | 2.33 | 6 | 0.0242 | 0.703 |
| Signaling by NOTCH1 t(7;9)(NOTCH1:M1580_K2555) Translocation Mutant | 74 | 7.19 | 13 | 0.0247 | 0.703 |
| Signaling by NOTCH1 in Cancer | 74 | 7.19 | 13 | 0.0247 | 0.703 |
| Signaling by NOTCH1 PEST Domain Mutants in Cancer | 74 | 7.19 | 13 | 0.0247 | 0.703 |
| FBXW7 Mutants and NOTCH1 in Cancer | 74 | 7.19 | 13 | 0.0247 | 0.703 |
| Signaling by NOTCH1 HD Domain Mutants in | 74 | 7.19 | 13 | 0.0247 | 0.703 |

| | | | | | |
|---|---|---|---|---|---|
| Cancer | | | | | |
| Signaling by NOTCH1 HD+PEST Domain Mutants in Cancer | 74 | 7.19 | 13 | 0.0247 | 0.703 |
| Signaling by NOTCH1 | 74 | 7.19 | 13 | 0.0247 | 0.703 |
| Synthesis of PC | 18 | 1.75 | 5 | 0.0251 | 0.703 |
| Formation of the active cofactor, UDP-glucuronate | 3 | 0.292 | 2 | 0.0265 | 0.728 |
| Mitotic G1-G1/S phases | 140 | 13.6 | 21 | 0.0287 | 0.729 |
| Autodegradation of Cdh1 by Cdh1:APC/C | 68 | 6.61 | 12 | 0.0293 | 0.729 |
| Nucleotide Excision Repair | 53 | 5.15 | 10 | 0.0294 | 0.729 |
| APC/C:Cdc20 mediated degradation of mitotic proteins | 76 | 7.39 | 13 | 0.0301 | 0.729 |
| Host Interactions of HIV factors | 141 | 13.7 | 21 | 0.0308 | 0.729 |
| Activated TLR4 signalling | 100 | 9.72 | 16 | 0.0309 | 0.729 |
| Sphingolipid de novo biosynthesis | 32 | 3.11 | 7 | 0.031 | 0.729 |
| Caspase-mediated cleavage of cytoskeletal proteins | 13 | 1.26 | 4 | 0.031 | 0.729 |
| Triglyceride Biosynthesis | 39 | 3.79 | 8 | 0.0312 | 0.729 |
| Stabilization of p53 | 54 | 5.25 | 10 | 0.033 | 0.743 |
| Activation of APC/C and APC/C:Cdc20 mediated degradation of mitotic proteins | 77 | 7.48 | 13 | 0.0332 | 0.743 |
| Signalling by NGF | 290 | 28.2 | 38 | 0.0334 | 0.743 |
| Coenzyme A biosynthesis | 8 | 0.778 | 3 | 0.0353 | 0.762 |
| Signaling by TGF-beta Receptor Complex | 70 | 6.8 | 12 | 0.0358 | 0.762 |
| GPVI-mediated activation cascade | 33 | 3.21 | 7 | 0.0361 | 0.762 |
| Antigen processing-Cross presentation | 78 | 7.58 | 13 | 0.0364 | 0.762 |
| Adaptive Immune System | 654 | 63.6 | 77 | 0.0386 | 0.771 |
| Toll Like Receptor 4 (TLR4) Cascade | 103 | 10 | 16 | 0.0393 | 0.771 |
| APC/C:Cdc20 mediated degradation of Securin | 71 | 6.9 | 12 | 0.0395 | 0.771 |
| Synthesis of DNA | 95 | 9.23 | 15 | 0.0397 | 0.771 |
| Cytokine Signaling in Immune system | 286 | 27.8 | 37 | 0.0419 | 0.771 |
| S Phase | 122 | 11.9 | 18 | 0.0469 | 0.771 |
| Cyclin E associated events during G1/S transition | 65 | 6.32 | 11 | 0.047 | 0.771 |
| Loss of proteins required for interphase microtubule organizationÃ‚Â from the centrosome | 65 | 6.32 | 11 | 0.047 | 0.771 |

| | | | | | |
|---|---|---|---|---|---|
| Loss of Nlp from mitotic centrosomes | 65 | 6.32 | 11 | 0.047 | 0.771 |
| MyD88:Mal cascade initiated on plasma membrane | 81 | 7.87 | 13 | 0.0475 | 0.771 |
| Toll Like Receptor TLR1:TLR2 Cascade | 81 | 7.87 | 13 | 0.0475 | 0.771 |
| Toll Like Receptor TLR6:TLR2 Cascade | 81 | 7.87 | 13 | 0.0475 | 0.771 |
| Toll Like Receptor 2 (TLR2) Cascade | 81 | 7.87 | 13 | 0.0475 | 0.771 |
| Hemostasis | 511 | 49.7 | 61 | 0.049 | 0.771 |
| Pentose phosphate pathway (hexose monophosphate shunt) | 9 | 0.875 | 3 | 0.0492 | 0.771 |
| Pyrimidine salvage reactions | 9 | 0.875 | 3 | 0.0492 | 0.771 |
| Regulation of cytoskeletal remodeling and cell spreading by IPP complex components | 9 | 0.875 | 3 | 0.0492 | 0.771 |
| NOTCH1 Intracellular Domain Regulates Transcription | 50 | 4.86 | 9 | 0.0492 | 0.771 |
| Assembly of the RAD50-MRE11-NBS1 complex at DNA double-strand breaks | 4 | 0.389 | 2 | 0.0496 | 0.771 |
| MRN complex relocalizes to nuclear foci | 4 | 0.389 | 2 | 0.0496 | 0.771 |
| Alpha-oxidation of phytanate | 4 | 0.389 | 2 | 0.0496 | 0.771 |

**Supplemental codes for reanalysis of single cell RNA sequencing data on R:**

**R code for analysis of GSE163668**

```
> library(Seurat)

> library(Matrix)

> library(dplyr)

>data_dir<- 'C:/Users/dclabiicb/Documents/GSE163668 pooled 5,6'

>expression_matrix<- Read10X(data.dir = data_dir)

>sample.tmp.seurat<- CreateSeuratObject(counts = expression_matrix, min.cells = 3, min.features = 200)

>sample.tmp.seurat<- NormalizeData(sample.tmp.seurat)

>sample.tmp.seurat<- FindVariableFeatures(sample.tmp.seurat, selection.method = "vst", nfeatures = 4000)

>nCoV.integrated<- ScaleData(sample.tmp.seurat)

>nCoV.integrated<- RunPCA(nCoV.integrated, verbose = FALSE,npcs = 100)

>nCoV.integrated<- ProjectDim(object = nCoV.integrated)

>nCoV.integrated<- FindNeighbors(object = nCoV.integrated, dims = 1:50)

>nCoV.integrated<- FindClusters(object = nCoV.integrated, resolution = 1.2)

>nCoV.integrated<- RunTSNE(object = nCoV.integrated, dims = 1:50)

>DimPlot(object = nCoV.integrated, reduction = 'tsne',label = TRUE)

>print(FeaturePlot(object = nCoV.integrated, features = c('PLAUR', 'ITGAX', 'HLA-DRA'),cols = c("lightgrey","Blue")))

>HLADRA_expression = GetAssayData(object = nCoV.integrated)["HLA-DRA",]

>ITGAX_expression = GetAssayData(object = nCoV.integrated)["ITGAX",]

>neg_ids = names(which(HLADRA_expression<0.5))

>neg_cells = subset(nCoV.integrated,cells=neg_ids)

>pos_ids = names(which(ITGAX_expression>0.5))

>pos_cells = subset(neg_cells,cells=pos_ids)

>FeaturePlot(pos_cells,"PLAUR", cols=c("lightgrey", "red"))

>neg_ids = names(which(HLADRA_expression<1))

>neg_cells = subset(nCoV.integrated,cells=neg_ids)

>pos_ids = names(which(ITGAX_expression>0.3))

>pos_cells = subset(neg_cells,cells=pos_ids)
```

```
>FeaturePlot(pos_cells,"PLAUR", cols=c("lightgrey", "red"))
```

**R code for analysis of GSE145926**

```
> library(Seurat)

> library(Matrix)

> library(dplyr)

> expression_matrix1 <- Read10X_h5(choose.files())

>sample.tmp.seurat1 <- CreateSeuratObject(counts = expression_matrix1, min.cells = 3,
min.features = 200)

> expression_matrix2 <- Read10X_h5(choose.files())

>sample.tmp.seurat2 <- CreateSeuratObject(counts = expression_matrix2, min.cells = 3,
min.features = 200)

>sample.tmp.seurat.combined<- merge(sample.tmp.seurat1, y = sample.tmp.seurat2)

> expression_matrix3 <- Read10X_h5(choose.files())

>sample.tmp.seurat3 <- CreateSeuratObject(counts = expression_matrix3, min.cells = 3,
min.features = 200)

>sample.tmp.seurat.combined.mild<- merge(sample.tmp.seurat.combined, y = sample.tmp.seurat3)

>sample.tmp.seurat.combined.mild<- NormalizeData(sample.tmp.seurat.combined.mild, verbose =
FALSE)

>sample.tmp.seurat.combined.mild<- FindVariableFeatures(sample.tmp.seurat.combined.mild,
selection.method = "vst", nfeatures = 2000,verbose = FALSE)

>nCoV.integrated<- ScaleData(sample.tmp.seurat.combined.mild, verbose = FALSE)

>nCoV.integrated<- RunPCA(nCoV.integrated, verbose = FALSE,npcs = 100)

>nCoV.integrated.mild<- ProjectDim(object = nCoV.integrated)

>nCoV.integrated<- FindNeighbors(object = nCoV.integrated, dims = 1:50)

>nCoV.integrated<- FindClusters(object = nCoV.integrated, resolution = 0.5)

>nCoV.integrated<- RunTSNE(object = nCoV.integrated, dims = 1:50)

>DimPlot(object = nCoV.integrated, reduction = 'tsne',label = TRUE)

>FeaturePlot(nCoV.integrated,features=c("ITGAX", "HLA-DRA", "PLAUR"), cols=c("light Grey",
"Blue"))

>VlnPlot(object = nCoV.integrated, features = c("ITGAX"),pt.size = 0)

>VlnPlot(object = nCoV.integrated, features = c("HLA-DRA"),pt.size = 0)

>neg_cells = subset(nCoV.integrated,cells=neg_ids)

>pos_ids = names(which(ITGAX_expression>0.5))
```

```
>pos_cells = subset(neg_cells,cells=pos_ids)
>FeaturePlot(pos_cells,"PLAUR", cols=c("light Grey", "Red"))
```

**R code for analysis of GSE168710**

```
>Raw.data<- readRDS(choose.files())
> metadata <- readRDS(choose.files())
> data <- CreateSeuratObject(Raw.data, meta.data = metadata, min.cells = 3, min.features = 200)
> data <- FindVariableFeatures(data, selection.method = "vst", nfeatures = 4000, verbose = FALSE)
> data <- ScaleData(data, verbose = FALSE)
>nCoV.integrated<- RunPCA(data, verbose = FALSE,npcs = 100)
>nCoV.integrated<- FindNeighbors(object = nCoV.integrated, dims = 1:50)
>nCoV.integrated<- FindClusters(object = nCoV.integrated, resolution = 1.2)
>nCoV.integrated<- RunTSNE(object = nCoV.integrated, dims = 1:50)
>DimPlot(object = nCoV.integrated, reduction = 'tsne',label = TRUE)
>DimPlot(object = nCoV.integrated, reduction = 'tsne',group.by= "stim",label = TRUE)
>FeaturePlot(nCoV.integrated,features=c("PLAUR"), cols=c("light Grey", "Blue"),label=TRUE)
>VlnPlot(object = nCoV.integrated, features = c("HLA-DRA"),pt.size = 0)
>VlnPlot(object = nCoV.integrated, features = c("ITGAX"),pt.size = 0)
>HLADRA_expression = GetAssayData(object = nCoV.integrated)["HLA-DRA",]
>ITGAX_expression = GetAssayData(object = nCoV.integrated)["ITGAX",]
>neg_ids = names(which(HLADRA_expression<2.5))
>neg_cells = subset(nCoV.integrated,cells=neg_ids)
>pos_ids = names(which(ITGAX_expression>0.8))
>pos_cells = subset(neg_cells,cells=pos_ids)
>FeaturePlot(pos_cells,features=c("PLAUR"), cols=c("light Grey", "Blue"))
>VlnPlot(pos_cells, features = c("PLAUR"),pt.size = 0)
>PLAUR_expression = GetAssayData(object = nCoV.integrated)["PLAUR",]
> PLAUR_2_ids = names(which(PLAUR_expression>2.2))
>  PLAUR_2_cells = subset(nCoV.integrated,cells=PLAUR_2_ids)
>DimPlot(object = PLAUR_2_cells, reduction = 'tsne',group.by= "stim",label = TRUE)
```
